# Bi-compartmental CSF–Serum Analysis of NfL and GFAP Differentiates Central and Peripheral Pathology in Neuroinfectious Diseases

**DOI:** 10.64898/2026.05.30.26354507

**Authors:** Deborah K. Erhart, Badrieh Fazeli, Franziska Bachhuber, Önder Soylu, Makbule Senel, Jan Lewerenz, Markus Otto, Steffen Halbgebauer, Hayrettin Tumani

## Abstract

**Background:** Neurofilament light chain (NfL) and glial fibrillary acidic protein (GFAP), established biomarkers of neuroaxonal injury and astroglial pathology, are frequently only assessed in blood, which limits conclusions regarding their origin. Bi-compartmental analyses of CSF and serum may help differentiate central or peripheral origin of biomarker elevation. Moreover, studies on NfL and GFAP in distinct neuroinfectious disease (NID) phenotypes are limited.

**Methods:** This retrospective monocentric study analyzed CSF and serum from patients with (meningo-)encephalitis/myelitis (TI+; *n*=48), meningitis (TI–; *n*=80), (cranial) nerve palsies/polyradiculitis (PND; *n*=61), and 113 non-neuroinflammatory/non-neurodegenerative controls. A bi-compartmental model using scatter plots and simple linear regression was applied to assess the origin of blood biomarker levels and discriminate between central and peripheral pathology.

**Results:** CSF and serum NfL and GFAP z-scores were significantly higher in TI+ compared with TI– (CSF-GFAP *p*<0.001/sGFAP *p*=0.0083; CSF-NfL *p*=0.003/sNfL *p*=0.0004). TI+ and PND differed only in GFAP levels, which were higher in TI+ (CSF-GFAP *p*=0.0049/sGFAP *p*=0.003). Bi-compartmental analysis revealed simultaneous elevation of CSF and serum NfL in TI+, indicating predominantly central origin, whereas PND demonstrated a shift toward higher sNfL levels suggesting peripheral origin. Higher clinical severity (modified Rankin Scale 3–5) was associated with elevated serum and CSF GFAP and NfL (sGFAP *p*=0.012/sNfL *p*=0.002; CSF-GFAP *p*<0.0001/CSF-NfL *p*=0.0001), which also predicted unfavorable outcome at discharge (sGFAP *p*=0.006/sNfL *p*=0.004; CSF-GFAP *p*=0.003/CSF-NfL *p*=0.012).

**Conclusions:** NfL and GFAP were associated with brain/myelon involvement in NID, predominantly reflecting central pathology. Despite strong CSF–serum correlations, bi-compartmental approaches provide additional insight into biomarker origin and disease compartment.

**What is already known on this topic:** Despite antiviral therapy, especially viral encephalitis remains associated with substantial mortality and long-term neurological sequelae. Neurofilament light chain (NfL) and glial fibrillary acidic protein (GFAP) are established biomarkers of neuroaxonal and astroglial injury and are increasingly used across neurological diseases. Previous studies in neuroinfectious diseases demonstrated elevated CSF and blood levels of NfL and GFAP, but mainly relied on central or peripheral compartments separately. These analyses do not fully reflect the origin of the measured biomarkers in either blood or CSF. Additionally, combined CSF–serum analyses with age-adjusted values in different manifestations of neuroinfectious diseases (NID), also in correlation with clinical outcome measurements, are limited.

**What this study adds:** This study demonstrates that NID patients with central nervous system involvement had higher CSF and serum NfL and GFAP values than those without. Bi-compartmental analyses using CSF–serum biomarker scatter plots and log– transformed values in a simple linear regression showed distinct patterns for encephalitis (bi-compartmental increase of values for NfL and GFAP), meningitis (similar to patient controls), and peripheral infectious nerve disease (NfL shift towards serum elevations). Biomarker values in CSF and serum at initial lumbar puncture (LP) were significantly higher in patients with longer hospitalization length and in patients with worse outcome at discharge according to the modified Rankin scale (mRS).

**How this study might affect research, practice or policy:** Our study offers a potential approach for the early biochemical detection of encephalitic involvement, possibly preceding overt clinical manifestations. Furthermore, as biomarker values at admission are associated with worse clinical outcome (mRS), those parameters could help clinicians in decision-making and in early detection of potential encephalitic courses.

## Introduction

Despite early antiviral therapy, mortality in herpes simplex virus type 1 (HSV-1) and varicella–zoster virus (VZV) encephalitis remains as high as 21.8% [1]. Survivors frequently develop complications or long-term sequelae, particularly cognitive decline (up to 70% in HSV-1 and 45% in VZV), markedly affecting daily functioning [2]. Current blood-based biomarker measurements do not reliably distinguish between central and peripheral sources of neuroaxonal and glial injury in neuroinfectious diseases. A combined CSF–serum (bi-compartmental) approach may overcome this limitation and improve pathophysiological interpretation.

Neurofilament light chain (NfL) and glial fibrillary acidic protein (GFAP), established indicators of structural central nervous system (CNS) damage in stroke, traumatic brain injury, multiple sclerosis, and neurodegeneration, increase following tissue damage [3–7]. Evidence in neuroinfectious disease (NID), particularly simultaneous CSF and blood measurements with longitudinal follow-up, remains limited. Elevated NfL and GFAP levels have been reported in herpesvirus infections (HSV-1, VZV), flavivirus-associated disease (tick-borne encephalitis (TBE), West Nile virus meningitis), and Lyme neuroborreliosis [8–14]. Notably, CSF and serum NfL are higher in VZV encephalitis than in Ramsay Hunt syndrome or meningitis,[11] and increased NfL in HSV-1 encephalitis is associated with poorer long-term outcomes [13]. However, the extent and origin of early tissue involvement remain unclear.

Ultrasensitive technologies such as single molecule array (Simoa) and microfluidic assays (Ella) enable reliable biomarker detection in serum [15]. The Ella platform demonstrates strong analytical performance and clinical potential, supported by age- adjusted reference values for NfL and GFAP, but has not yet been applied in NID [16–20].

In our study, we aimed to (I) detect differences in CSF and serum NfL and GFAP in various manifestations of a disease, i.e. with or without brain tissue involvement or peripheral nerve disease, (II) investigate the correlation of CSF and serum NfL and GFAP with routine CSF findings, clinical findings, and outcome parameter (modified Rankin scale; mRS), (III) build a bi-compartmental pattern of NfL to identify the origin of the elevated values (i.e. centrally or peripherally), and (IV) detect temporal dynamics of serum and CSF levels of NfL and GFAP in follow-up analyses.

## Methods

### Study design, participants and selection process

In our retrospective cohort study, we included 189 patients with NID (encephalitis, meningitis, myelitis, cranial nerve palsies, polyradiculitis, or a combination thereof according to clinical case definitions [14, 21–24]) from the Department of Neurology (University Hospital of Ulm, Germany) based on their clinical, radiographic, and CSF findings (see further information in the Supplementary Material). CSF findings (leukocyte count, QAlb, total protein, oligoclonal IgG-bands, CSF-C-X-C motif chemokine ligand 13 (CSF-CXCL13)) and demographical data (age, sex, admission to ICU/IMC, time since onset to LP, hospitalization length, mRS at admission/discharge) were taken from the medical charts. Additionally, we included 116 patient controls (PC) with no clinical, radiographic, or CSF findings suggestive of neuroinflammatory or neurodegenerative (CNS) disease. The patient selection process, additional information on included NID aetiologies, and diseases of the PC cohort are illustrated in Figure 1, Tables S1+S2 (Supplementary Material).

**Figure 1.**
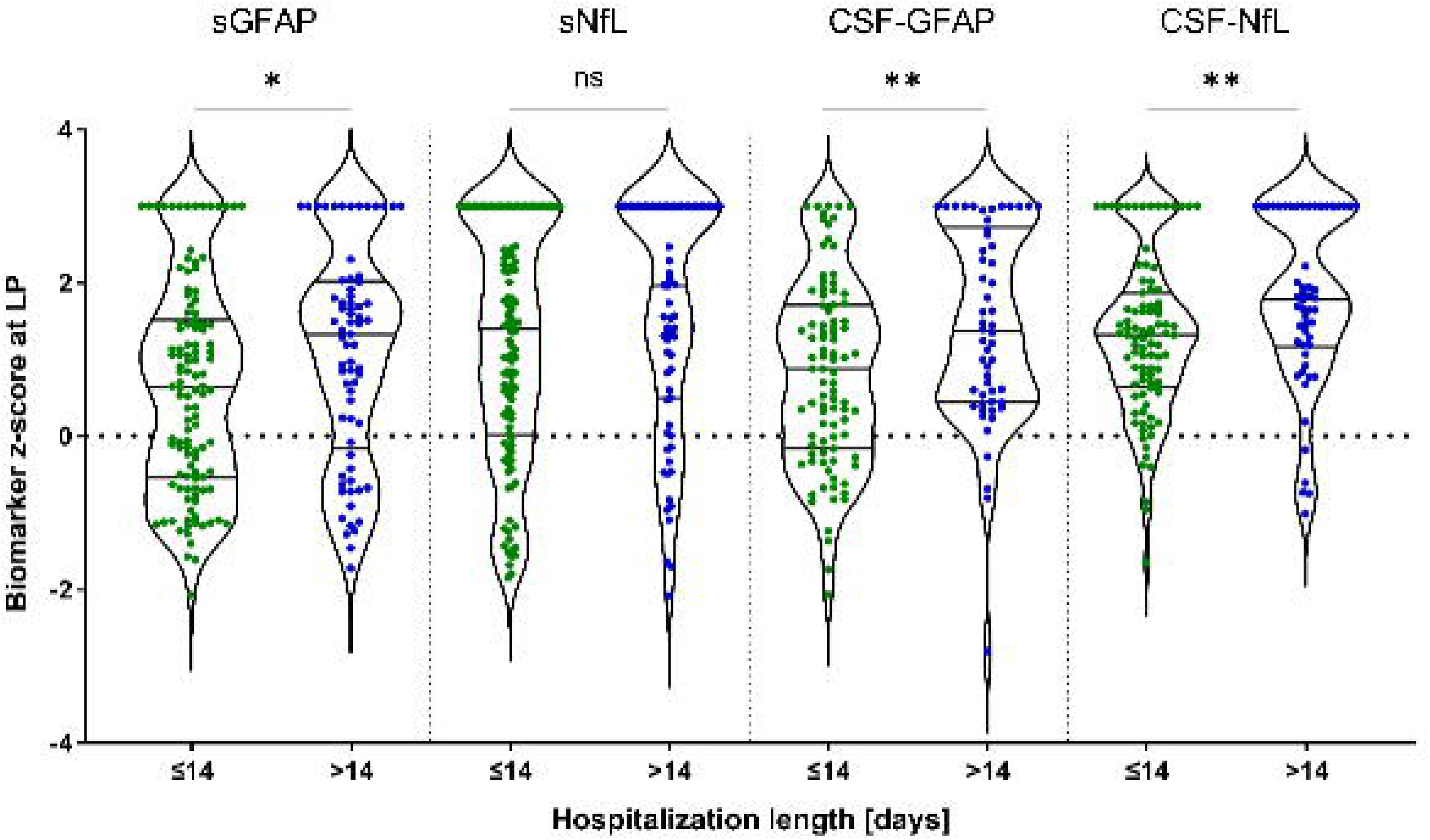
Patient selection process. The patient selection process is described in this flowchart. Due to suspected neuroinfectious disease (NID), 451 patients of the biobank of the Department of Neurology, University Hospital of Ulm, Germany, received a diagnostic lumbar puncture from August 2009 to November 2023. According to the above criteria, 189 patients were selected for further measurements of glial fibrillary acidic protein (GFAP) and neurofilament light chains (NfL) in cerebrospinal fluid (CSF) and serum. CSF: cerebrospinal fluid, TI+: NID with tissue involvement, TI–: NID without tissue involvement, PND: peripheral nerve disease.

### CSF and biomarker analyses

CSF and blood samples were obtained during the diagnostic workup. All patients were treatment-naïve with regard to antibiotic and antiviral therapy. Further CSF laboratory analyses and direct and indirect pathogen detection is described in the Supplementary Material.

GFAP and NfL levels were quantified in CSF (CSF-GFAP, CSF-NfL) and serum (sGFAP, sNfL) using commercially available Ella assay kits (Bio-Techne, Minneapolis, USA). Measurements of CSF-CXCL13 were performed using the ELISA from Quantikine R&D Systems (Minneapolis, MN, USA) and Euroimmun (Lübeck, Germany; method comparison test R^2^=0.97). All measurements were performed according to each manufacturer’s instructions. For further information, see Supplementary Material.

### Ethics

All patients gave their written informed consent before lumbar- and venipuncture. The study is in accordance with the Declaration of Helsinki and the STROBE guidelines and was approved by the local ethics committee of the University of Ulm (ethics approval number 20/10; 3^rd^ May 2010).

### Statistical analysis

Group comparisons with more than two groups were analyzed non-parametrically due to the non-Gaussian distribution of the data (Shapiro-Wilk test). Continuous variables were compared using the Kruskal-Wallis test with Dunn’s post hoc test, categorical variables with Pearson’s χ² test (SPSS, V.28.0.1.0 (142), IBM Corp., Armonk, NY, USA), and pairwise comparisons with the Mann-Whitney U test.= Continuous data are presented as median (IQR: 25^th^−75^th^ percentile), categorical data as counts and percentages.

NfL and GFAP concentrations were normalized to age-adjusted z-scores [17, 18]. Since no age-reference curve exists for CSF-GFAP, z-scores were derived from the age-matched PC cohort using the formula: z=(X−μ_controls_)/σ_controls_ with log10-transformed values, mean (μ_controls_), and standard deviation (σ_controls_). All z-scores >2.5 were set to 3.0. Data visualization and additional analyses, including Spearman correlations (incl. 95% confidence interval (CI) of the Spearman’s rank coefficient *r*), were performed in GraphPad Prism (V.10.2.2 (GraphPad Software, La Jolla, CA)). CSF–serum scatter plots were generated in R (V.4.5.2 (R Foundation, Vienna, Austria)) using log–transformed values and simple linear regression with ±3 SD prediction intervals. Statistical significance was set at *p*<0.05. Missing data were handled by pairwise exclusion.

## Results

### Demographic, clinical, and routine CSF findings

A total of 189 patients were assigned to three clinically defined groups (TI+, TI–, PND; Table S1; Supplementary Material). TI– comprised 42.3% (*n*=80) with meningitis with/without peripheral nerve involvement, while 48 (25.4%) and 61 (32.3%) patients were classified as TI+ and PND, respectively (Table S2; Supplementary Material). In TI+, 89.5% had (meningo-)encephalitis; in PND, 80.3% had neuritis cranialis. An overview of the demographic, clinical, and diagnostic parameters can be found in Table 1 (whole NID cohort with available serum samples) and Tables S3+S4 (Supplementary Material). Routine CSF parameters and raw CSF NfL and GFAP values differed between groups and pathogens. Intergroup differences are summarized in Table 1, and Tables S3+S5 (Supplementary Material). The median age of the PC cohort, which was solely used for z-standardization of CSF-GFAP, was 50.0 (IQR, 23.25–61.0) years. The sex distribution 46.55% male (*n*=54) and 53.45% female (*n*=62).

**Table 1.** Demographical characteristics and CSF parameters of the three groups with available serum samples.

| **Demographics/**<br>**CSF parameters** | **TI+**<br>**($n=47$)** | **TI–**<br>**($n=80$)** | **PND**<br>**($n=60$)** | **P-value** |
| --- | --- | --- | --- | --- |
| *n*<br>(female/<br>male) | 31/16<br>(65.96%/<br>34.04%) | 51/29<br>(63.7%/<br>36.3%) | 41/19<br>(68.33%/<br>31.67%) | 0.814<sup>*</sup> |
| Age [years] | 66.0 (55.5–<br>76.0) | 41.0 (31.25–<br>57.5) | 57.0 (39.0–<br>68.0) | <0.001<sup>**</sup> |
| Time since onset<br>to LP [days] | 6.0 (2.0–21.0) | 4.0 (2.0–8.75) | 4.0 (1.0–12.5) | 0.122<sup>**</sup> |
| sGFAP [pg/ml] | 12.7 (6.6–<br>41.2) | 4.3 (2.2–9.0) | 5.1 (3.1–9.2) | <0.0001<sup>**</sup> |
| sNfL [pg/ml] | 67.6 (23.4–<br>206) | 13.93 (8.5–<br>33.5) | 54.0 (26.6–<br>155) | <0.0001<sup>**</sup> |
| Duration of<br>hospitalization<br>[days] | 18.0 (14.0–<br>23.25) | 13.0 (7.0–<br>15.0) | 14.0 (8.0–<br>15.0) | <0.001<sup>**</sup> |
| Admission to<br>ICU/IMC; [+%], *n* | 100%<br>(47/0) | 1.3%<br>(1/79) | 0%<br>(0/60) | <0.001<sup>*</sup> |
| (+/-) |  |  |  |  |
| Leucocyte count<br>[/μl] | 72.5 (22.0–<br>158.25) | 148.5 (63.5–<br>317.75) | 83.0 (31.0–<br>184.5) | **<0.001\*\*** |
| Lactate [mmol/l] | 2.19 (1.72–<br>2.9) | 2.0 (1.8–2.6) | 2.0 (1.7–2.49) | 0.441** |
| Total protein [mg/l] | 960.0 (576.5–<br>1726.25) | 724.0 (530.0–<br>1018.5) | 861.0 (562.5–<br>1348.0) | **0.043\*\*** |
| QAlb (CSF/serum<br>albumin) | 14.4 (9.0–<br>25.2) | 9.5 (7.2–<br>15.28) | 12.2 (7.6–<br>21.8) | **0.021\*\*** |
| CSF-CXCL13<br>[pg/ml] | 211.5 (37.0–<br>465.25) | 91.0 (28.25–<br>259.75) | 1170.5 (45.75–<br>13450.0) | **0.002\*\*** |
| CSF-specific<br>oligoclonal IgG<br>bands; [+%], *n* (+/-<br>) | 46.81%<br>(22/25) | 28.75%<br>(23/57) | 53.33%<br>(32/28) | **0.025\*** |

Group differences among TI+, TI–, and PND were assessed using Pearson's $\chi^2$ test\* for categorical variables (sex, CSF-specific oligoclonal IgG bands) and the Kruskal–Wallis test\*\* for all other continuous variables. Categorical data are presented as counts and percentages, and continuous data as median and interquartile range (IQR, 25<sup>th</sup>–75<sup>th</sup> percentile). Statistical significance was set at $p < 0.05$ . Significant values are highlighted in bold. TI+: NID with tissue involvement, TI–: NID without tissue involvement, PND: peripheral nerve disease, CSF: cerebrospinal fluid, LP: lumbar puncture, ICU/IMC: intensive care unit, intermediate care unit, QAlb: CSF-albumin/serum-albumin, CXCL13: C-X-C-motif chemokine ligand 13.

### GFAP and NfL values in different cohorts

Z-score values for each biomarker are presented by groups in Figure 2. All four biomarkers demonstrated significantly higher z-scores in the TI+ group compared to the TI− group (sGFAP *p*=0.0083; sNfL *p*<0.0001; CSF-NfL *p*=0.0003; CSF-GFAP *p*<0.0001), with the most pronounced difference observed in CSF-GFAP and sNfL. When comparing the TI+ and PND groups, GFAP z-scores showed significant differences in both serum and CSF (sGFAP *p*=0.0030; CSF-GFAP *p*=0.0049), whereas no significant difference was observed for sNfL (*p*>0.9999) and CSF-NfL (*p*=0.2452). In the comparison between the TI− and PND groups, significantly elevated z-score levels of sNfL (*p*<0.0001) and CSF-GFAP (*p*=0.0435) were observed in the PND group. Absolute biomarker values, including comparisons to the PC cohort, are presented in Figure S1 (Supplementary Material).

**Figure 2.**
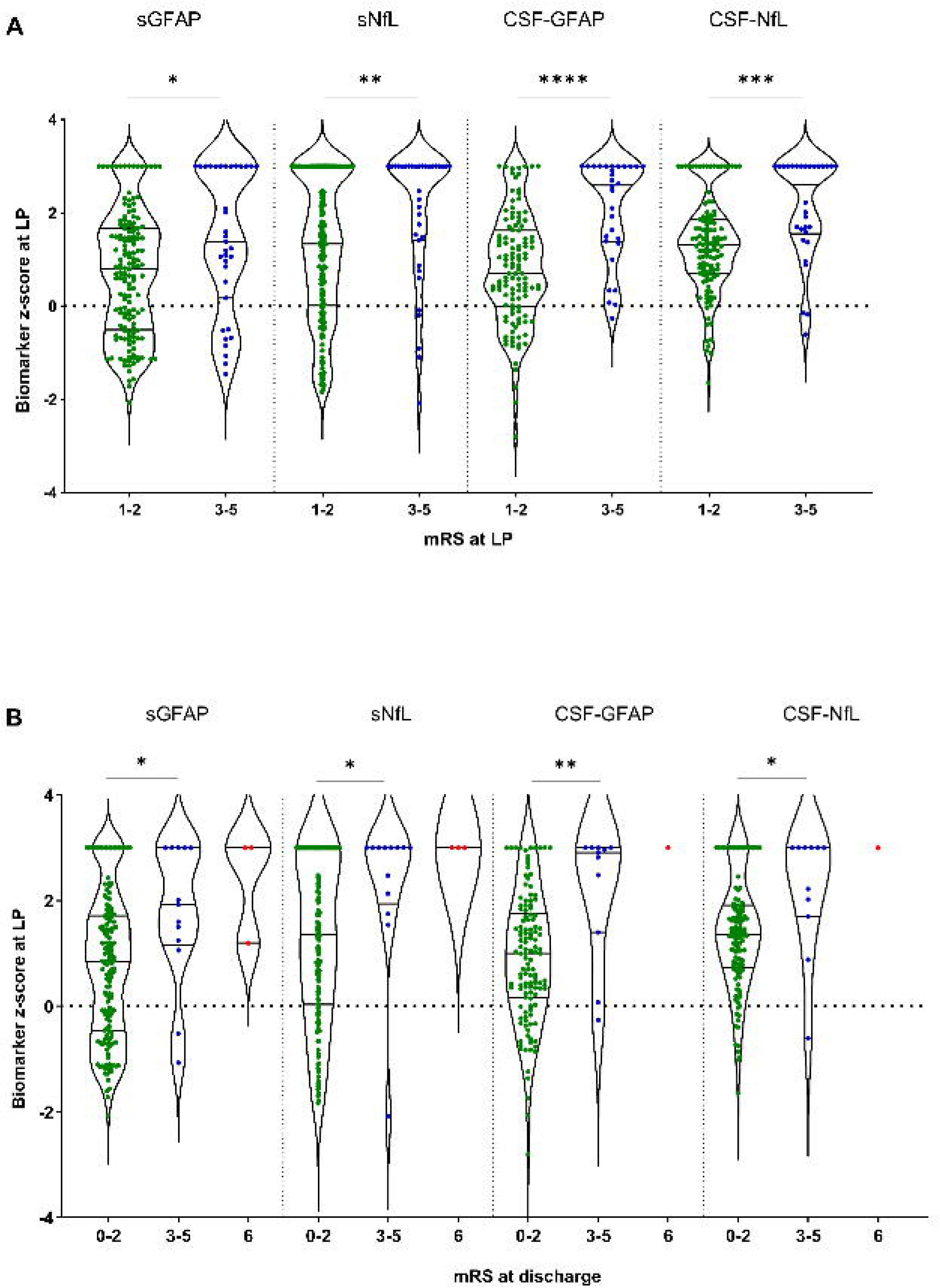
Z-scores for serum and CSF biomarkers across disease groups. Violin plots show the distribution of age-corrected z-scores for sGFAP and sNfL (Panels A and B) and CSF-NfL and CSF-GFAP (Panels C and D) across three clinical groups: NID with tissue involvement (TI+), NID without tissue involvement (TI−), and peripheral nerve disease (PND). (A) sGFAP z-scores were significantly elevated in the TI+ group compared to both TI− and PND groups (**p<0.01). (B) sNfL z-scores were significantly higher in the TI+ and PND group compared to TI− (****p<0.0001). (C) CSF-GFAP z-scores showed significant differences between all three groups, with the highest values in the TI+ group (*p<0.05, **p<0.01, ****p<0.0001). (D) CSF-NfL z-scores were significantly elevated in the TI+ group compared to TI− (***p<0.001). Black dots represent individual subjects; thick horizontal bars indicate median values; violin widths reflect data distribution. Dotted line indicates a z-score of 0.

### Correlation between GFAP and NfL in serum and CSF

Spearman correlations between serum and CSF biomarkers were analyzed separately by group (Figure S2; Supplementary Material). The strongest and most consistent association was observed between sNfL and CSF-NfL across all groups (*r*=0.73–0.79; all *p*<0.0001). In contrast, GFAP correlations were group-dependent: sGFAP and CSF-GFAP showed moderate correlations in PC (*r*=0.60 (95%CI, 0.42–0.73)), TI– (*r*=0.48 (95%CI, 0.25–0.66)), and PND (*r*=0.49 (95%CI, 0.23–0.69)) but a strong correlation in TI+ (*r*=0.76 (95%CI, 0.58–0.86); all *p*≤0.0005). Correlations between sGFAP and sNfL were moderate in PC (*r*=0.53 (95%CI, 0.38–0.66)), TI–(*r*=0.57 (95%CI, 0.40–0.71)), and PND (*r*=0.53 (95%CI, 0.31–0.70), all *p*<0.0001) and weaker in TI+ (*r*=0.44 (95%CI, 0.16–0.65); *p*=0.002). CSF-GFAP and CSF-NfL correlations ranged from moderate to strong, lowest in TI+ (*r*=0.50 (95%CI, 0.23–0.70), *p*=0.001) and highest in PND (*r*=0.70 (95%CI, 0.50–0.82), *p*<0.0001).

Serum GFAP correlated with CSF-CXCL13 only in the PND cohort (*r*=0.32 (95%CI, 0.006–0.59), *p*=0.04). In contrast, sNfL showed a strong correlation with CSF-CXCL13 in TI+ (*r*=0.67 (95%CI, 0.39–0.84), *p*<0.0001). CSF-GFAP did not correlate with CSF-CXCL13 in any cohort, whereas CSF-NfL correlated significantly in TI+ (*r*=0.50 (95%CI, 0.12–0.75), *p*=0.0113). Further correlation analyses between serum and CSF values of NfL and GFAP and routine CSF findings in the different NID cohorts revealed several significant associations which are fully displayed in Figure S3 (Supplementary Material), Tables S6+S7 (Supplementary Material).

### Bi-compartmental analysis

To explore the correlation between biomarker concentrations in serum and CSF, values for each biomarker were plotted on log–log scatter plots (Figure 3 and Figure S4, Supplementary Material). To enable comparison with normal patterns observed in control individuals, we focused on NfL, for which we recently analyzed a large control cohort and published a model defining the normal reference range for serum and CSF concentrations [18]. In the present study, the same model was applied, and patient values were overlaid onto this reference framework. As shown, internal control patients (grey) fell within the established control range (Figure 3). In contrast, patients in the TI+ group showed a clear increase in both serum and CSF NfL concentrations. This pattern was not observed in the TI− group, whose values largely overlapped with those of the control population. In the PND group, serum NfL concentrations were markedly elevated compared with controls.

**Figure 3.**
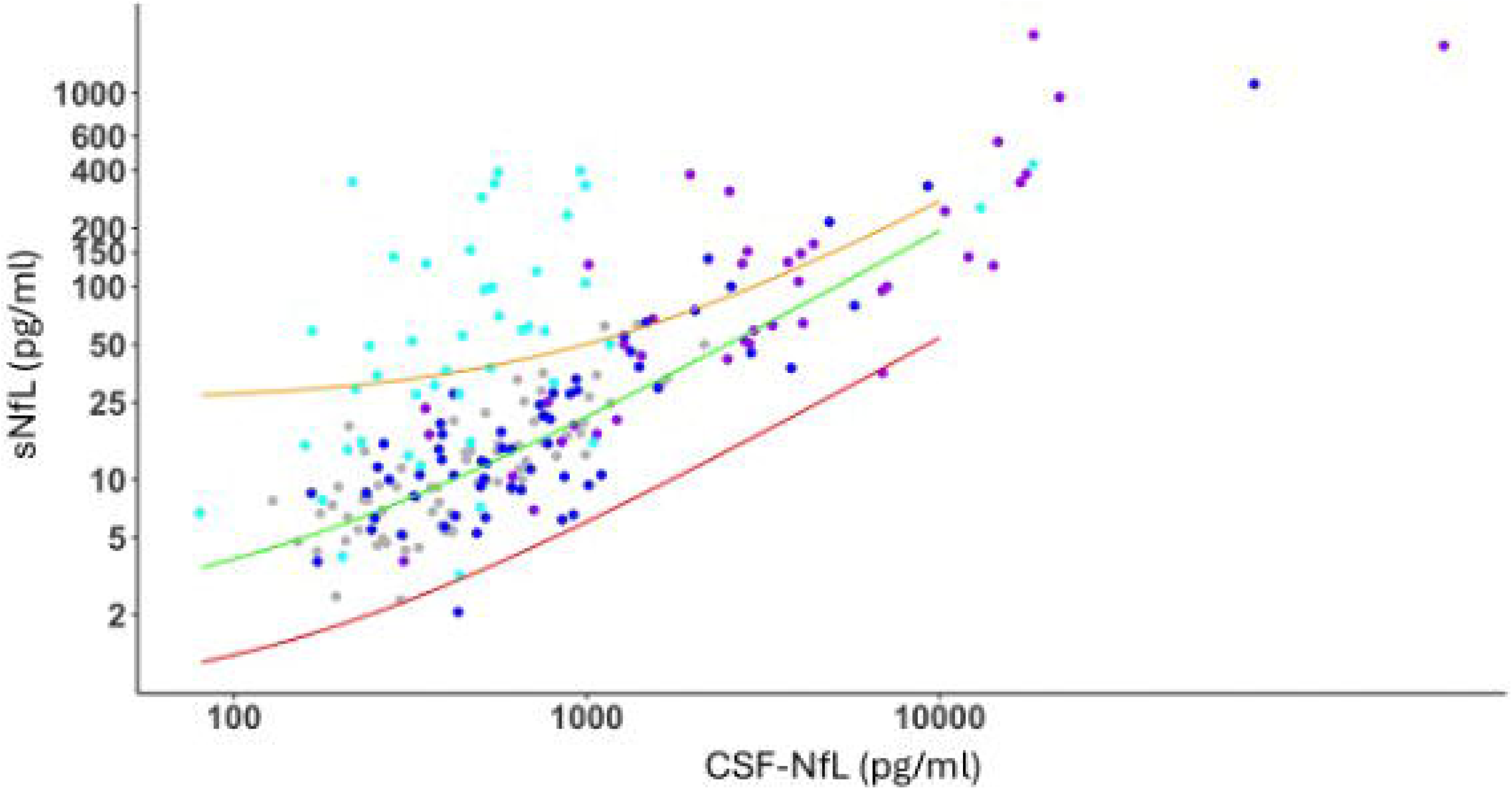
Relationship between CSF and serum NfL concentrations across study groups. Scatter plot showing CSF versus serum NfL concentrations. Axes are plotted on a logarithmic scale. Each point represents an individual participant, color-coded by group (internal control group: grey; TI+: purple; TI−: blue; PND: cyan). Solid lines represent linear regression models fitted to log-transformed values, with corresponding prediction intervals shown in red (lower) and orange (upper).

As no sufficiently large control cohort was available to establish a comparable reference model for GFAP, we used our internal PC cohort and these data were plotted together with the TI+, TI− and PND groups on log–log scatter plots and are presented in Figure S4 (Supplementary Material). For GFAP, both serum and CSF concentrations are clearly increased in the TI+ group, whereas values in the other two groups largely remain within the range observed in the PC cohort.

### Associations of GFAP and NfL levels with clinical outcome

In a subsequent analysis, we examined whether the modified Rankin scale (mRS) score at the time of initial LP was associated with the measured biomarkers NfL and GFAP in both CSF and serum (Figure 4A). The entire NID cohort was stratified into two groups according to clinical severity (mRS_low_: 1–2 vs. mRS_high_: 3–5) as described previously [25]. Serum GFAP (*p*=0.012) and NfL levels (*p*=0.002) were significantly elevated in the mRS_high_ cohort. This pattern could be also demonstrated and was pronounced in the corresponding CSF measurements (CSF-GFAP: *p*<0.0001; CSF-NfL: *p*=0.0001). We further assessed whether biomarker levels at initial LP were associated with clinical outcome as reflected by mRS at discharge (Figure 4B). In the overall NID cohort, three patients (all from the TI+ subgroup) died and were classified as non-survivors. A favorable outcome (mRS 0–2) occurred in 85.2% of the patients. Serum and CSF concentrations of GFAP and NfL at initial LP were significantly higher in patients with an unfavorable outcome (mRS 3–5 at discharge) compared to those with a favorable outcome (mRS 0–2) (Figure 4B; sGFAP: *p*=0.006, sNfL: *p*=0.004, CSF-GFAP: *p*=0.003, CSF-NfL: *p*=0.012).

**Figure 4.**
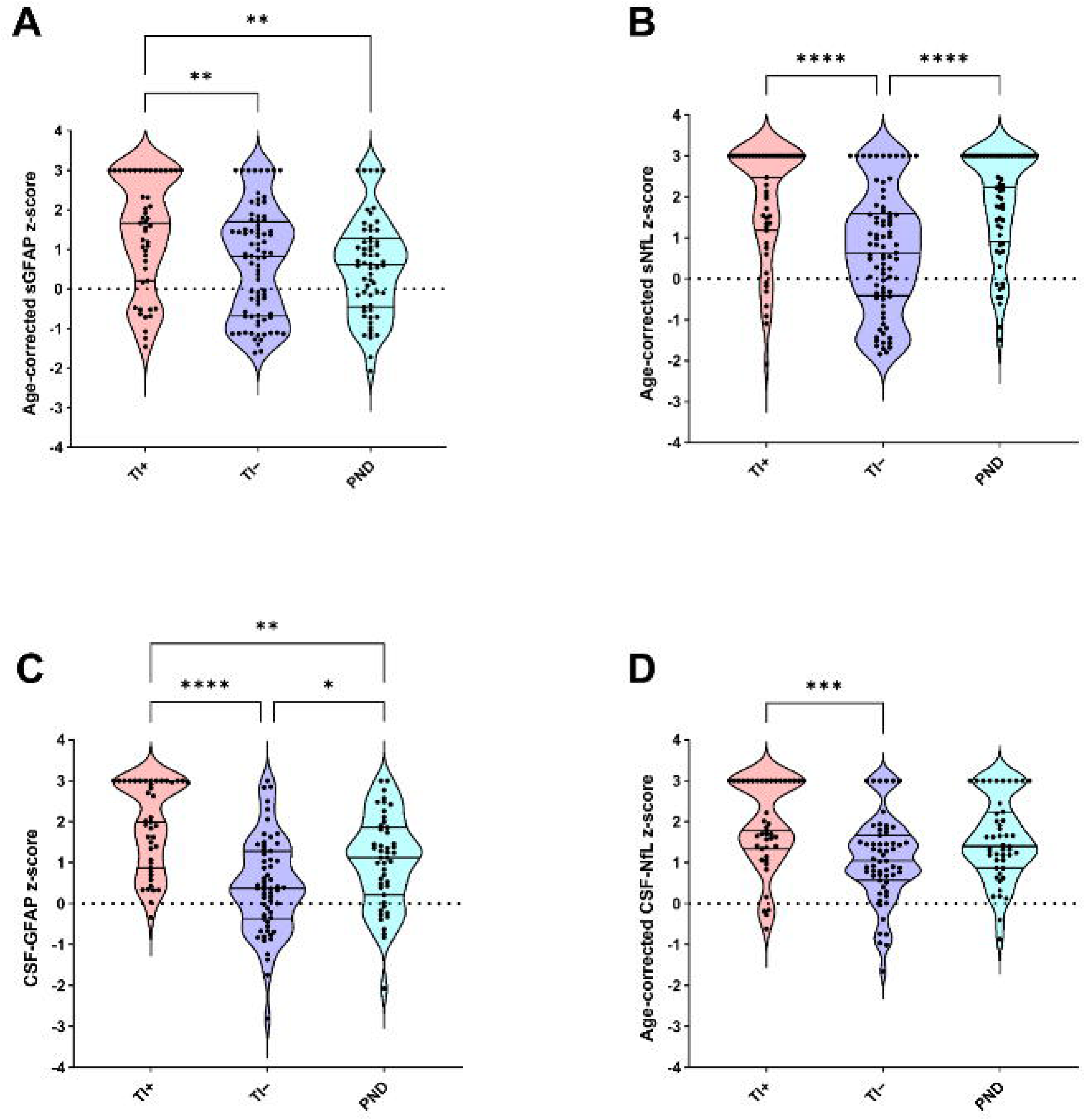
Associations between biomarker levels and modified Rankin Scale (mRS) at first LP and at discharge. Violin plots show the distribution of age-corrected z-scores for CSF-GFAP, sGFAP, CSF-NfL, and sNfL for the whole neuroinfectious disease (NID) cohort according to mRS_low_ (1-2) vs. mRS_high_ (3-5) at first LP (A) and at discharge (B). Differences between the groups were calculated using Mann-Whitney U test. Statistical significance was set at p<0.05 (*p<0.05, **p<0.01, ***p=0.0001, ****p<0.0001). Black dots represent individual subjects; horizontal lines within the violin plots indicate median values and IQR (25th−75th percentile); violin widths reflect data distribution. Dotted line indicates a z-score of 0.

Furthermore, the whole NID cohort was stratified by its median hospitalization length of 14 days (≤14 vs. >14 days; Figure 5). Patients with elevated biomarker levels, except sNfL (*p*=0.092), exhibited significantly longer hospital stays compared to those with lower concentrations (sGFAP: *p*=0.004; CSF-GFAP: *p*=0.036, CSF-NfL: *p*=0.004). This association was observed for both serum and CSF measurements, with particularly pronounced differences in the CSF-based stratification (Figure 5).

**Figure 5.**
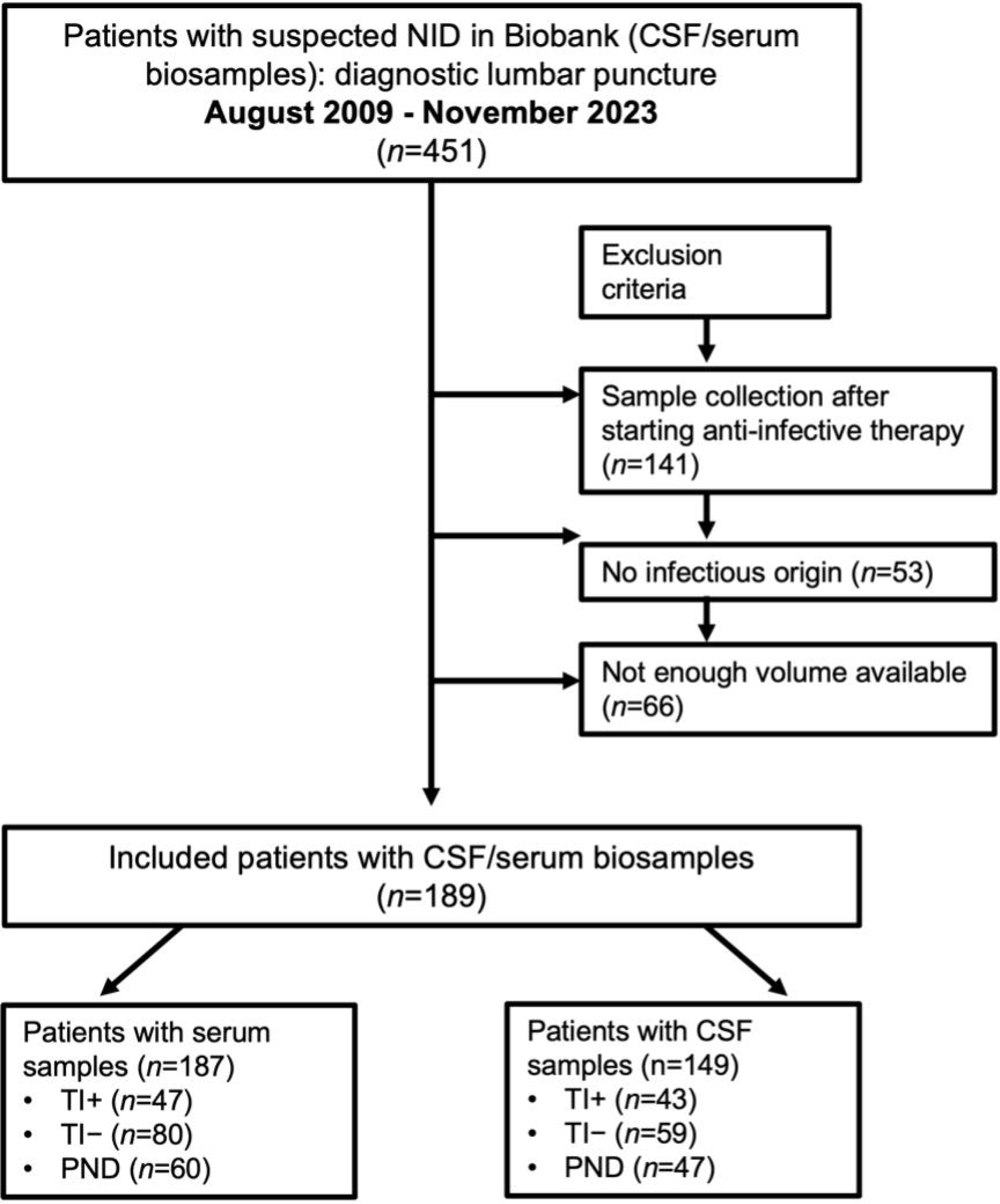
Associations of different biomarker levels after cohort stratification according to hospitalization length (days). Violin plots show the differences in serum and CSF NfL and GFAP z-score values after stratification of the whole NID cohort according to the calculated median of hospitalization in days (14 days). Pairwise comparisons were calculated using the Mann-Whitney U test. P-values <0.05 were set as significant. *p<0.05, **p<0.01, ns: not significant.

### Follow-up serum and CSF analyses of GFAP and NfL

Follow-up lumbar punctures within 4 weeks were available for 57 patients (TI+: 10; TI–: 33; PND: 14). Compared with PC (sGFAP median 3.2 (IQR, 1.7–6.6) pg/ml, sNfL median 12.2 (IQR, 7.7–19.3) pg/ml, CSF-GFAP median 248 (IQR, 172–358) pg/ml, CSF-NfL median 416 (IQR, 259–670) pg/ml), serum and CSF NfL and GFAP at baseline were markedly elevated in TI+ (all *p*<0.0001; Figure S5; Supplementary Material). During follow-up (TI+ cohort), CSF-GFAP remained significantly elevated after 1 week (median 1180 (IQR, 1080–1268) pg/ml, *p*=0.0360), CSF-NfL after 2 (median 1592 (IQR, 1112–13926) pg/ml, *p*=0.0148) and 3 weeks (median 2916 (IQR, 1128–93548) pg/ml, *p*=0.0406), and sNfL after 3 weeks (median 134 (IQR, 59.4– 1178) pg/ml, *p*=0.0412).

In TI– (Figure S6; Supplementary Material), neither serum nor CSF NfL or GFAP showed relevant longitudinal changes. In contrast, the PND cohort (Figure S7; Supplementary Material) demonstrated elevated baseline NfL in serum (*p*=0.0096) and CSF (*p*=0.0278), but not GFAP (sGFAP: *p*>0.9999, CSF-GFAP: *p*=0.1426); sNfL remained significantly increased at weeks 1 (*p*=0.0114) and 2 (*p*=0.0042) versus PC.

## Discussion

We conducted a comprehensive study assessing NfL and GFAP levels in CSF and serum across a large, heterogeneous cohort of neuroinfectious diseases, including longitudinal measurements using the microfluidic Ella platform and age-corrected z-scores. NfL (type IV) and GFAP (type III) intermediate filament proteins are established markers of neuroaxonal damage and astroglial pathology [3, 26]. Due to significant intergroup age differences and their known age dependency [18, 27, 28], biomarker levels were analyzed as absolute values and age-corrected z-scores, with group comparisons primarily based on z-scores.

In TI+ patients, all four biomarkers showed significantly elevated z-scores compared with TI– consistent with previous reports of higher GFAP and NfL in encephalitis than meningitis [8, 9, 29]. Evidence on GFAP in neuroinfectious disease is limited, but increasing data highlight astrocyte activation (increase in number and change of phenotype) proportional to CNS inflammation severity [8, 9, 29, 30]. Correlation analyses aligned with previous findings [9, 31], demonstrating strong CSF–serum NfL associations across groups, while GFAP correlations were strongest in TI+ again underscoring the severity of CNS inflammation. Neuroaxonal or astroglial injury leads to release of NfL and GFAP into the CSF, with subsequent passage into the bloodstream depending on blood–CSF barrier integrity [3, 26].

Both biomarkers reflect distinct pathological processes, as indicated by their differential concentration patterns. No significant difference in CSF-NfL levels was observed between the TI– and PND cohorts, whereas sNfL levels differed significantly between these groups. To further investigate this finding, we applied a preliminary bi-compartmental analytical approach, which to date has been established only for NfL [18]. Given the moderate to strong association between CSF and serum NfL concentrations, this method enabled the identification and graphical illustration of the likely origin of blood NfL elevations [18]. In TI+ patients, the CSF/serum NfL ratio was simultaneously shifted toward higher values in both compartments compared with the TI– and PND groups, suggesting a predominantly central origin of serum NfL. This pattern likely reflects a higher degree of direct neurotoxic intrathecal inflammation, as observed, for example, in necrotizing and frequently hemorrhagic herpes simplex encephalitis (HSE) [13]. HSV-1 exerts direct cytotoxic and lytic effects on neurons, often leading to neuronal destruction and hemorrhagic transformation [32]. In contrast, the proposed pathophysiology of VZV, also an α-herpesvirus with 64 of 70 genes homologous to HSV-1, is primarily characterized by vasculopathy affecting small- to large-caliber cerebral arteries and the vasa vasorum of peripheral cranial nerves, frequently resulting in polyneuritis cranialis [32]. Neurotoxic effects are thought to occur secondarily through ischemic transaxonal processes with subsequent astrogliosis and axonal injury [32]. Nevertheless, a small cohort study comparing CSF GFAP and NfL levels in VZV and HSV-1 encephalitis reported no significant differences [14], whereas another small cohort study found significant differences in CSF NfL and GFAP between HSE and TBE patients [29]. However, only one patient in the TBE group had encephalitis [29]. Consistently, MRI evidence of extensive invasive neuroinflammation was observed in our study exclusively in the TI+ cohort, including mesiotemporal lesions (18.8%), basal ganglia or thalamic involvement (10.4%), multifocal T2-weighted and contrast-enhancing white matter hyperintensities (14.6%), and brainstem lesions (10.4%). In contrast, the PND group demonstrated predominantly elevated NfL levels in blood relative to the other cohorts, in agreement with previous findings [18]. This observation is consistent with the clinical composition of the PND cohort, which comprised patients with peripheral infectious, predominantly cranial nerve, involvement. Our data indicate a predominantly peripheral origin of blood NfL elevations attributable to axonal damage. In contrast, the TI– cohort showed a slight shift toward higher CSF-NfL concentrations, consistent with the raw data demonstrating significantly increased CSF-NfL levels compared with PC. This observation is supported by studies of chemokine profiles distinguishing HSV-1 encephalitis from HSV-2 and VZV meningitis, which reported pronounced systemic inflammation in encephalitis characterized by markedly elevated CXCL8, CXCL9, and CXCL10 levels in both CSF and serum, whereas meningitis was associated with predominantly intrathecal production of CXCL11 and CCL8 [33, 34]. Although there is a substantial clinical demand for blood-based biomarkers of neuronal injury, the present findings demonstrate that bi-compartmental analyses are essential for determining the origin of biomarkers detected in blood [18]. Despite the strong correlation between CSF and serum concentrations of NfL and GFAP, blood measurements may reflect neuroaxonal damage or glial pathology arising from either central or peripheral neuroinfectious processes [11, 18].

Additionally, significant correlations with CSF-CXCL13 were observed. This marker is strongly elevated in early neuroborreliosis, even before pleocytosis or total protein increases, and has been proposed as an indicator of disease severity [35, 36]. In encephalitis, CSF-CXCL13 correlates with serum and CSF NfL, reflecting intrathecal inflammation with direct neuronal and axonal injury [3, 35]. Accordingly, CXCL13-driven B-cell recruitment may be contribute to parenchymal CNS damage and increased NfL release. In contrast, meningitis and infectious cranial nerve palsies mainly affect the meninges or peripheral nerves, with limited CNS axonal injury, potentially explaining the absent or weak correlations between NfL and CSF-CXCL13 in these conditions. Notably, GFAP showed no significant correlation with CSF-CXCL13.

The temporal dynamics of these biomarkers in neuroinfectious diseases have been only sparsely studied [13, 14, 29]. Available evidence suggests that biomarker concentrations vary over time. For NfL in HSE, peak levels occur approximately 8–14 days after disease onset, followed by a gradual decline, although values remain substantially elevated compared with patient controls [13, 29]. In contrast, GFAP concentrations appear highest during the acute phase (<7 days) and subsequently decline steeply toward near-normal levels over several weeks to months [13, 29]. Another follow-up study examining both biomarkers in a clinically heterogeneous cohort of VZV infections (*n*=16) demonstrated a slight increase in CSF NfL and GFAP levels after 10–15 days, followed by a decrease after 3–5 months [14]. In the present study, longitudinal analyses were limited by the small number of available serial biosamples. Nevertheless, sNfL and CSF-NfL values were still significantly elevated for up to 3 weeks in the TI+ and PND cohort. One possible explanation, particularly in HSE, might be the persistence of a proinflammatory state, with soluble markers such as neopterin, interleukin-2 receptor, and cluster of differentiation 8 remaining elevated for several weeks beyond the acute phase [37, 38]. In addition, extensive neuroinflammation may induce Wallerian anterograde degeneration, leading to disruption of myelin sheaths, destruction of the axonal cytoskeleton, and involvement of proximal neuronal segments and cell bodies, as previously described in Japanese B encephalitis [39].

To date, only a limited number of studies have examined the prognostic significance of NfL and GFAP in neuroinfectious diseases. NfL has been suggested to predict long-term neurocognitive outcome in patients with HSE, potentially in the context of secondary anti-NMDA receptor (N-methyl-D-aspartate receptor) autoimmunization, as CSF-NfL levels were shown to strongly correlate with anti-NMDA receptor IgG titers [13]. In contrast, a cohort study of thirteen patients with VZV encephalitis found no significant association between CSF concentrations of NfL or GFAP and cognitive outcome after a median follow-up of 41.5 months (range 19–85 months) [12]. In the present study, we evaluated short-term functional outcome as measured by mRS at discharge. The mRS has been widely used to describe functional outcome after any disease and especially neuroinfectious diseases [2]. Elevated biomarker levels at the time of initial lumbar puncture were associated with higher mRS scores at discharge and prolonged hospitalization, both in serum and CSF. These findings support the potential utility of NfL and GFAP as prognostic biomarkers in neuroinfectious diseases.

This study has several strengths, including a well-characterized cohort of NID patients with simultaneous CSF and serum sampling, integration of clinical severity and outcome measures, and the application of a novel bi-compartmental analytical approach enabling inference on biomarker origin. The use of age-adjusted z-scores further enhances comparability across individuals.

However, several limitations should be considered. The retrospective, monocentric design may limit generalizability. The heterogeneous spectrum of pathogens and clinical presentations introduces potential variability, and classification based on the predominant phenotype may not fully capture mixed disease manifestations. In addition, incomplete CSF–serum pairing and the absence of an external validation cohort for the bi-compartmental model restrict broader applicability. Finally, the lack of established reference standards for CSF-GFAP limits direct comparability with NfL-based analyses.

Taken together, we demonstrate that combined CSF-serum assessment of NfL and GFAP provides clinically meaningful insights into the extent and anatomical origin of neuroaxonal and astroglial injury in neuroinfectious diseases. Bi-compartmental analysis reveals distinct biomarker patterns that differentiate central versus peripheral involvement, thereby extending beyond the interpretive limitations of single-compartment measurements. Furthermore, bi-compartmental biomarker assessment might offer a potential approach for the early biochemical detection of encephalitic involvement, possibly preceding overt clinical manifestations. Future prospective studies should therefore investigate patients initially presenting with meningitic disease courses who subsequently develop encephalitis, to determine whether changes in CSF–serum biomarker relationships can serve as early indicators of central nervous system involvement. Given the currently limited data, multicenter studies are warranted to validate this framework and to explore its potential integration into routine diagnostic workflows.

## Data Availability

All data produced in the present study are available upon reasonable request to the authors from qualified investigators.

## Acknowledgement

We want to thank Refika Aksamija, Joleene Holm, Vera Lehmensik, Nicole Renske, Tatiana Simak, Kevin Liese, Sandra Hübsch, Dagmar Schattauer, and Alice Beer from the CSF laboratory/Biobank of the neurological department of the University of Ulm for technical assistance, preparation, and providing of the biosamples.

