## Supplementary Material for "Bi-compartmental CSF–Serum Analysis of NfL and GFAP Differentiates Central and Peripheral Pathology in Neuroinfectious Diseases"

[†]Shared-first authorship

[‡]Shared-last authorship

*Corresponding authors:

Deborah K. Erhart:

Hayrettin Tumani:

### Supplementary Material

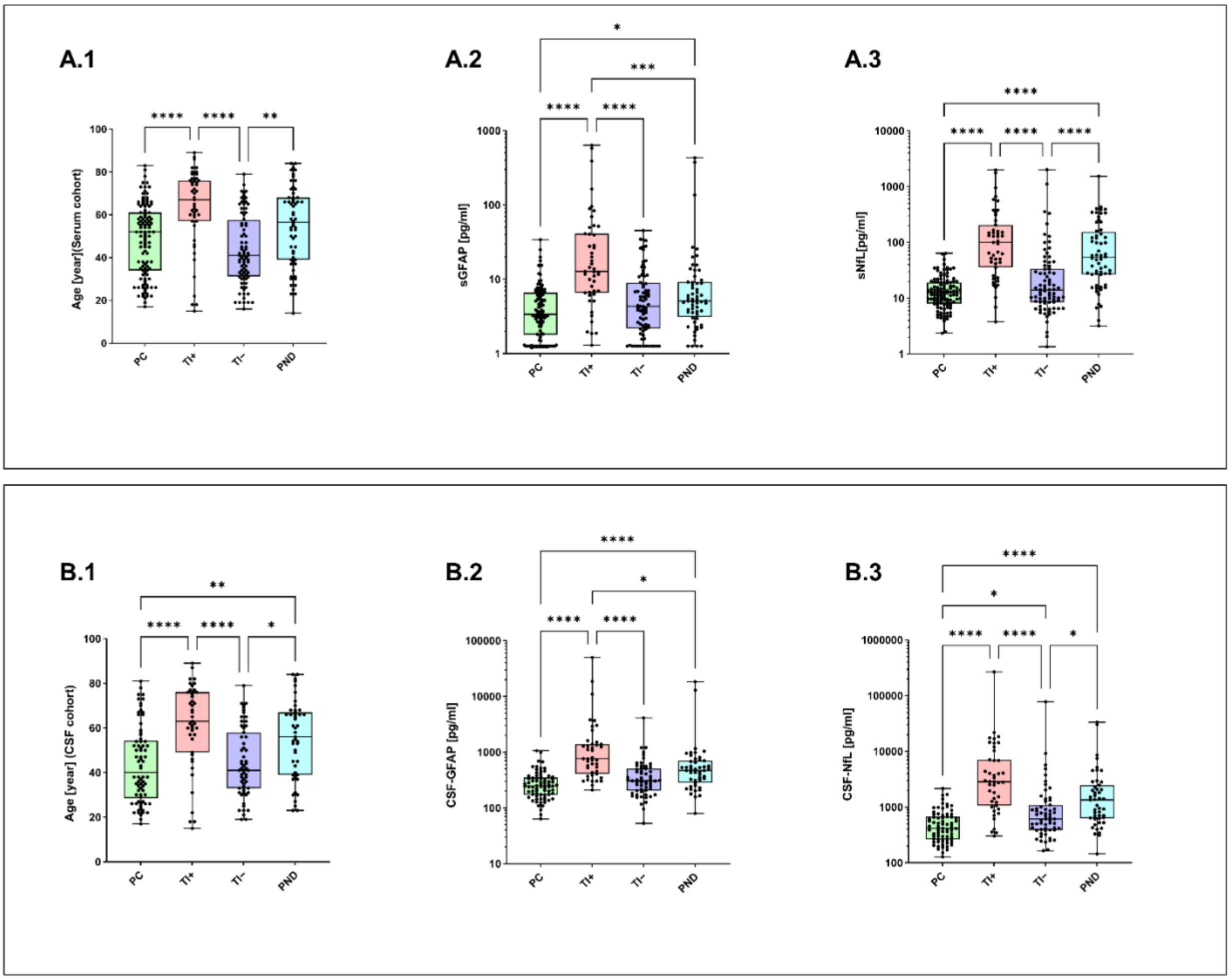


**Figure S1. CSF and serum absolute values of GFAP and NfL in the clinical cohort.**

Box plots illustrate (A.1) the age distribution in the serum cohort and (B.1) the age distribution in the CSF cohort; (A.2) serum GFAP concentrations; (A.3) sNfL concentrations; (B.2) CSF-GFAP concentrations; and (B.3) CSF-NfL concentrations across four clinical groups: patient controls (PC), neuroinflammatory disease with tissue involvement (TI+), neuroinflammatory disease without tissue involvement (TI−), and peripheral nerve disease (PND). Data are presented as median with interquartile range (IQR, 25th–75th percentile). Biomarker concentrations are displayed on a logarithmic y-axis. Group differences were assessed using the Kruskal–Wallis test followed by Dunn's post hoc analysis (*$p$<0.05; **$p$<0.01; ***$p$<0.001; ****$p$<0.0001). Statistical significance was set at $p$-value <0.05.

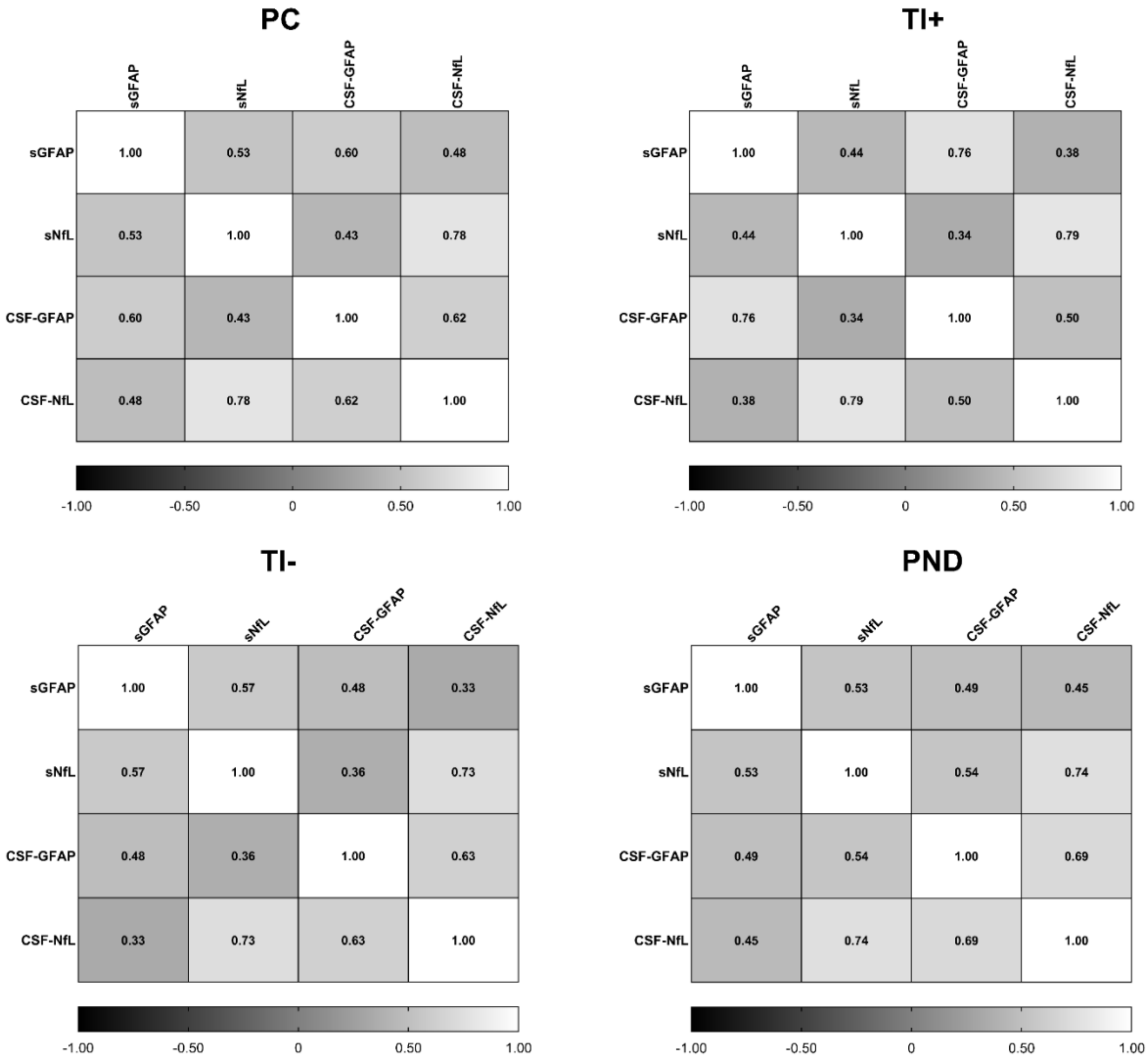


**Figure S2. Correlation matrices of serum and CSF biomarkers across study groups.**

Heatmaps illustrate pairwise Spearman correlation between serum and CSF levels of GFAP and NfL within each group: PC (patient controls), TI+ (NID with tissue involvement), TI– (NID without tissue involvement), and PND (peripheral nerve disease). Each heatmap shows correlation coefficients (*r*-values), with color intensity indicating the strength and direction of correlation (from -1 to +1).

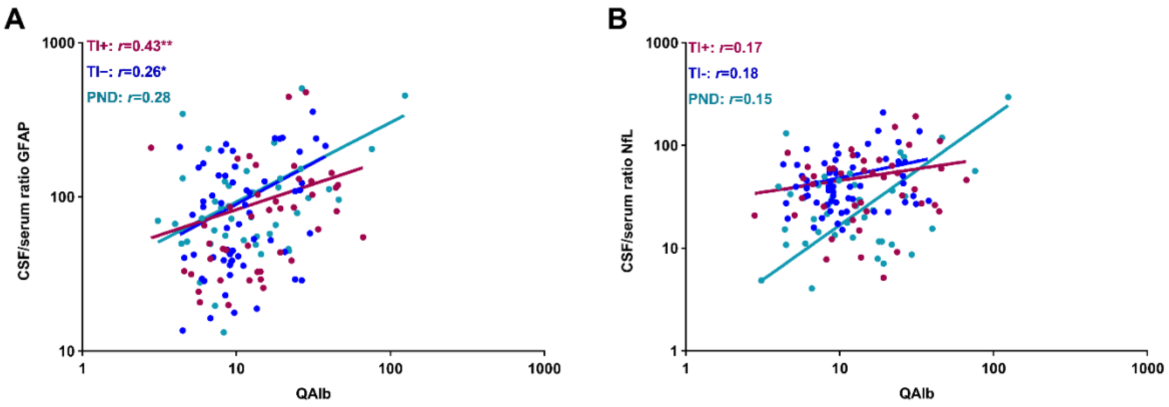


**Figure S3. Correlations between QAlb and CSF/serum GFAP and NfL ratios.**
Scatterplots illustrate the Spearman correlation between the albumin quotient (QAlb) and the CSF/serum ratios of GFAP (A) and NfL (B) across three patient cohorts: TI+ (NID with tissue involvement, pink), TI– (NID without tissue involvement, blue), and PND (peripheral nerve disease, cyan). Each line represents the regression trend for the respective cohort. Spearman's correlation coefficients (*r*) are indicated in the figure. Axes are plotted on a logarithmic scale. Asterisks denote levels of significance (**p*<0.05; ***p*<0.01).

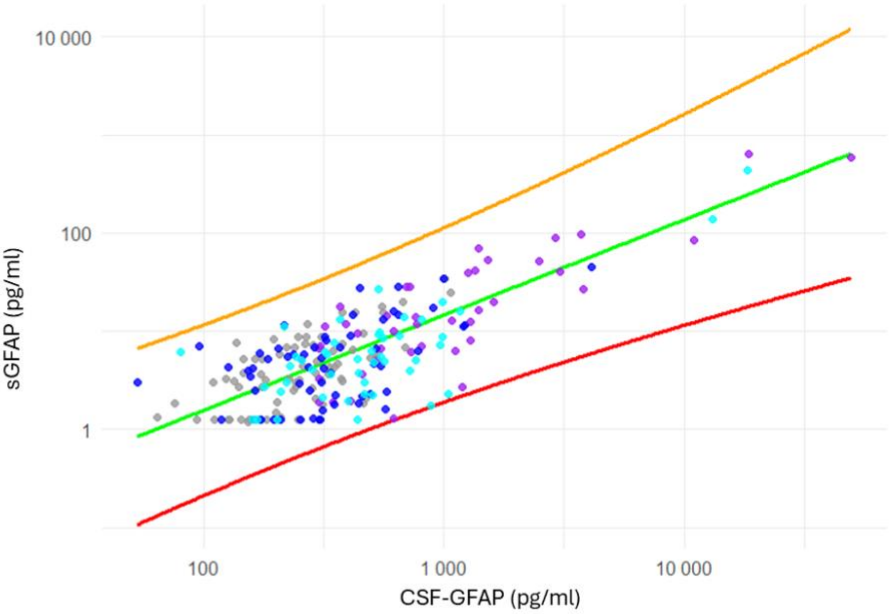


**Figure S4. Association between serum and CSF GFAP concentrations in a bi-compartmental pattern.**
Serum and CSF GFAP concentrations were log10-transformed and plotted on linear axes. Linear regression was performed on log-transformed data. Each point represents an individual patient, color-coded by group (PC: grey; TI+: purple; TI−: blue; PND: cyan). The TI+ cohort exhibited overall consistently elevated GFAP levels in both serum and CSF, suggesting systemic involvement of inflammation. In contrast, the PND and TI− cohorts showed GFAP levels comparable to those of the PC cohort according to this bicompartmental pattern.

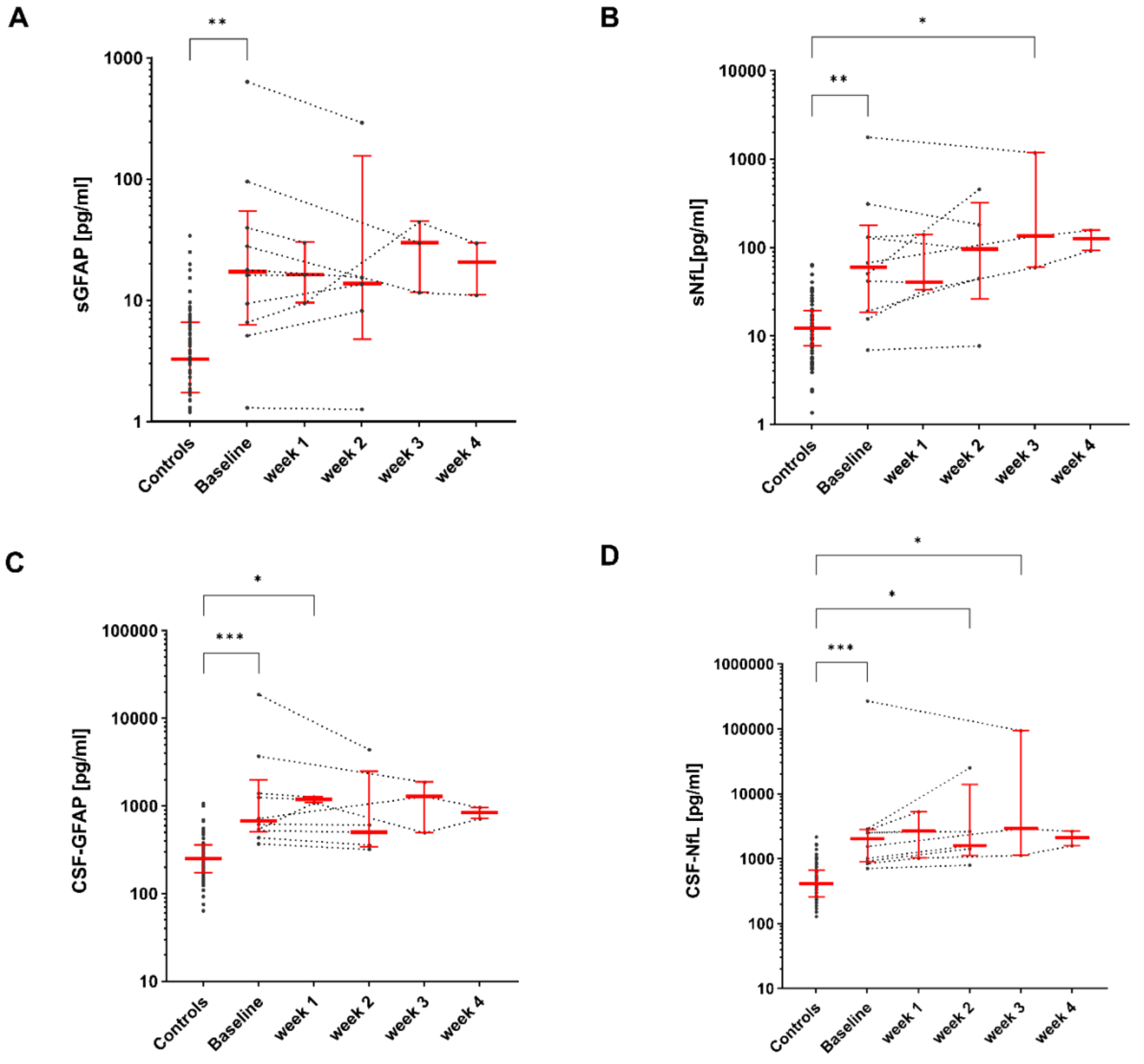


**Figure S5. Follow-up analyses of serum and CSF GFAP and NfL in the TI+ cohort.**

Scatter plot illustrates baseline and follow-up analyses of serum and CSF values of GFAP and NfL in the TI+ (tissue involvement+) cohort up to 4 weeks. Values for the control group and individual participants are given as median and IQR (25$^{th}$–75$^{th}$ percentile). The y-axis is displayed on a logarithmic scale. Groups were compared by Kruskal-Wallis test and Dunn's post hoc test (**p*<0.05; ***p*<0.01; ****p*<0.001; *****p*<0.0001). *P*-values <0.05 were regarded as statistically significant.

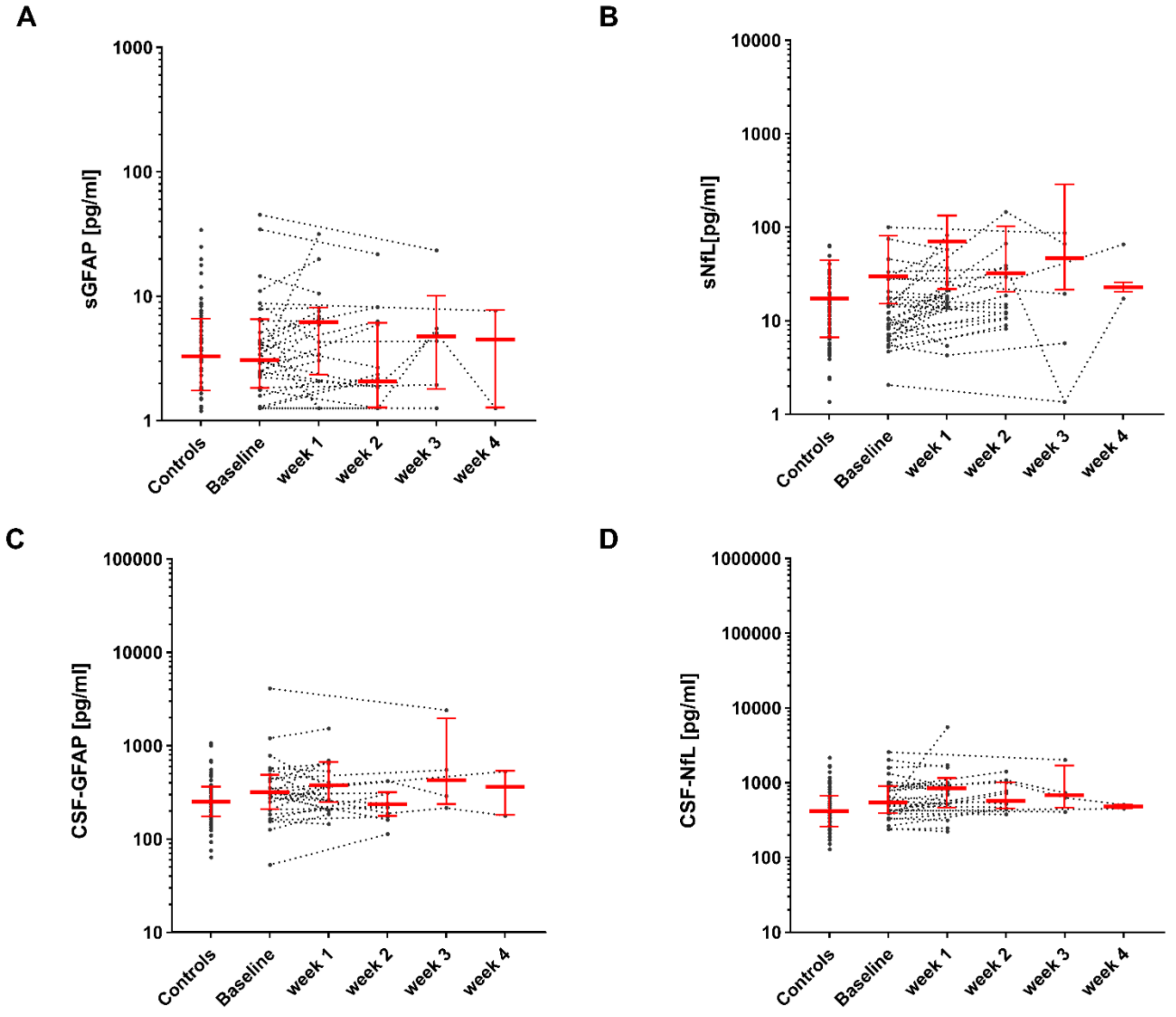


**Figure S6. Follow-up analyses of serum and CSF GFAP and NfL in the TI– cohort.**

Scatter plot illustrates baseline and follow-up analyses of serum and CSF values of GFAP and NfL in the TI– (tissue involvement–) cohort up to 4 weeks. Values for the control group and individual participants are given as median and IQR ($25^{th}$–$75^{th}$ percentile). The y-axis is displayed on a logarithmic scale. Groups were compared by Kruskal-Wallis test and Dunn's post hoc test (**p*<0.05; ***p*<0.01; ****p*<0.001; *****p*<0.0001). *P*-values <0.05 were regarded as statistically significant. CSF-GFAP/sGFAP: CSF/serum glial fibrillary acidic protein, CSF-NL/sNfL: CSF/serum neurofilament light chain.

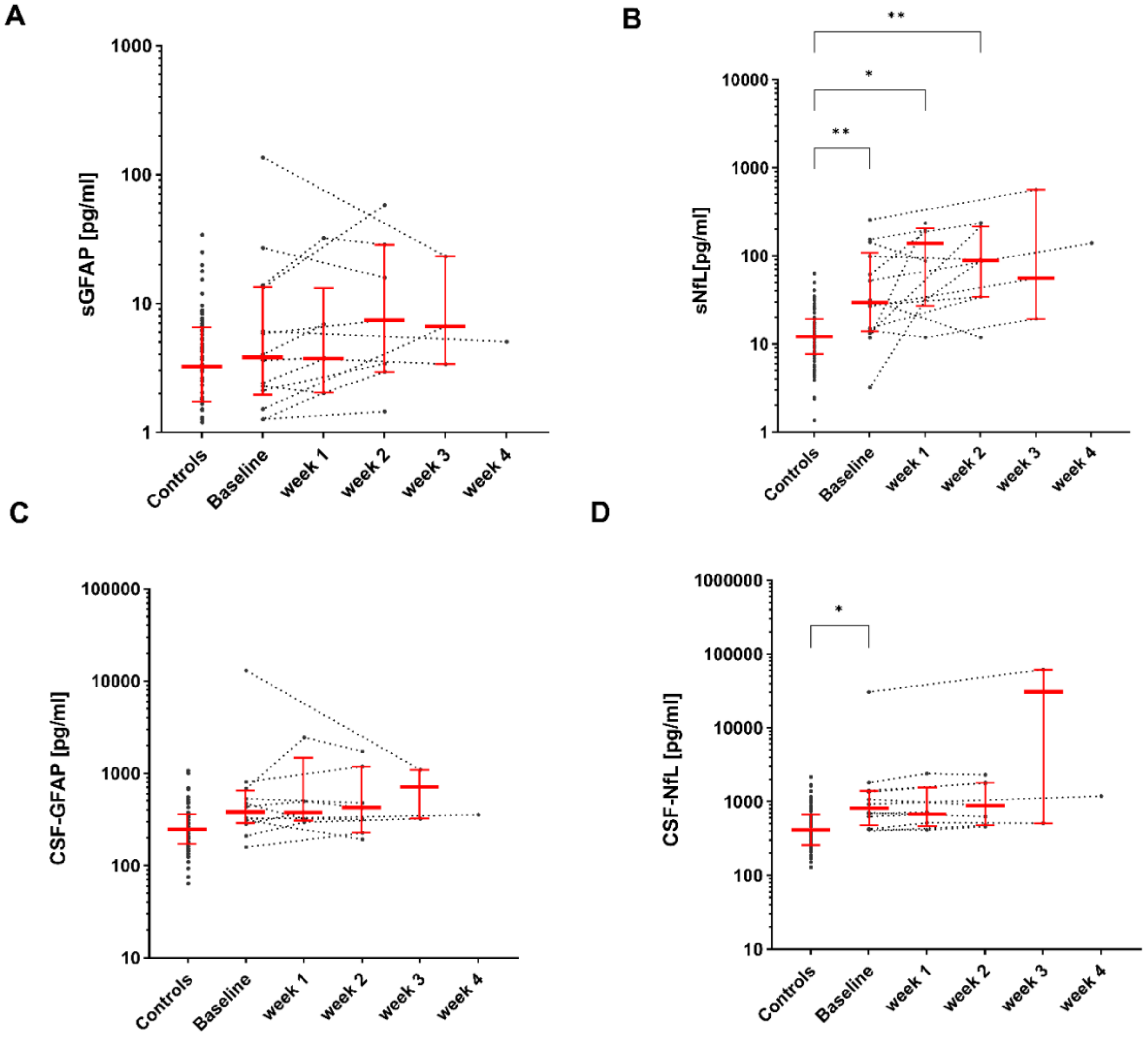


**Figure S7. Follow-up analyses of serum and CSF GFAP and NfL in the PND cohort.**

Scatter plot illustrates baseline and follow-up analyses of serum and CSF values of GFAP and NfL in the PND (peripheral nerve disease) cohort up to 4 weeks. Values for the control group and individual participants are given as median and IQR (25$^{th}$–75$^{th}$ percentile). The y-axis is displayed on a logarithmic scale. Groups were compared by Kruskal-Wallis test and Dunn's post hoc test (**p*<0.05; ***p*<0.01; ****p*<0.001; *****p*<0.0001). *P*-values <0.05 were regarded as statistically significant. CSF-GFAP/sGFAP: CSF/serum glial fibrillary acidic protein, CSF-NL/sNfL: CSF/serum neurofilament light chain.

**Table S1. Case definitions and diagnostic criteria [1-5].**

| |
|---|
| **Encephalitis**<br>• *Major criteria (both required):*<br> • Encephalopathy ≧24 hours<br> • No other explanation detectable<br>• *Minor criteria (at least two):*<br> • Objective fever (≧38°C)<br> • CSF analysis: White blood cell count ≧5/µl<br> • New onset of focal neurological deficits<br> • New onset seizures<br> • EEG findings consistent with neurological dysfunction<br> • Brain MRI findings consistent with encephalitis |
| **Meningitis**<br>• New onset of headache, light sensitivity, nausea/vomiting<br>• Objective fever (≧38°C)<br>• No signs of parenchymatous involvement<br>• CSF analysis: White blood cell count ≧5/µl<br>• CSF culture/antigen/PCR suggestive for infectious cause (except aseptic form) |
| **Myelitis**<br>• MRI findings consistent with myelopathy<br>• CSF analysis: White blood cell count ≧5/µl<br>• Clinical findings suggestive for spinal cord involvement (paresis, autonomic dysfunction regarding bladder/bowel, sensibility dysfunction) |
| **Cranial nerve palsy (infectious cause)**<br>• Peripheral nerve palsies of one or more cranial nerves<br>• CSF analysis: White blood cell count ≧5/µl |
| **Polyradiculitis (infectious cause)**<br>• Inflammation of the spinal nerve roots, accompanied by weakness, sensory loss/paresthesia, pain<br>• CSF analysis: White blood cell count ≧5/µl |

CSF: cerebrospinal fluid, EEG: electroencephalogram, MRI: magnetic resonance imaging, PCR: polymerase chain reaction.

**Table S2. Pathogens and related diagnoses in the three different groups.**

| **Pathogen** | **TI+ (*n*=48; 25.4%)** | **TI– (*n*=80; 42.3%)** | **PND (*n*=61; 32.3%)** |
|---|---|---|---|
| Tick-borne encephalitis virus (*n*=12; 6.3%) | • Meningoencephalitis (*n*=2) | • Meningitis (*n*=10) | |
| Herpes simplex virus type 1 (*n*=9; 4.85%) | • (Meningo-) Encephalitis (*n*=7) | | • (Poly-) Neuritis cranialis (*n*=1)<br>• Polyradiculitis (*n*=1) |
| Herpes simplex virus type 2 (*n*=2; 1.1%) | | • Meningitis (*n*=2) | |
| Varizella zoster virus (*n*=50; 26.5%) | • (Meningo-) Encephalitis (*n*=11) | • Meningitis (*n*=11)<br>• Meningitis + (poly-) neuritis cranialis (*n*=4)<br>• Meningitis + polyradiculitis (*n*=2) | • (Poly-) Neuritis cranialis (*n*=21)<br>• Polyradiculitis (*n*=1) |
| Epstein-Barr virus (*n*=1; 0.5%) | | • Meningitis (*n*=1) | |
| Hepatitis E virus (*n*=1; 0.5%) | • (Meningo-) Encephalitis (*n*=1) | | |
| John Cunningham virus (*n*=1; 0.5%) | • Encephalitis (*n*=1) | | |
| Staphylococcus aureus/epidermidis/Streptococcus anginosus/Citrobacter freundii (*n*=4; 2.1%) | • (Meningo-) Encephalitis (*n*=2)<br>• Abscess (*n*=1) | • Meningitis + polyradiculitis (*n*=1) | |
| Borrelia burgdorferi (*n*=37; 19.6%) | • (Encephalo-) Myelitis (*n*=3)<br>• Myelitis (*n*=1)<br>• Myelitis + (poly-) neuritis cranialis (*n*=2) | • Meningitis (*n*=1)<br>• Meningitis + (poly-) neuritis cranialis (*n*=2)<br>• Meningitis + polyradiculitis (*n*=1) | • Polyradiculitis (*n*=7)<br>• (Poly-) Neuritis cranialis (*n*=18)<br>• Polyradiculitis + (poly-) neuritis cranialis (*n*=2) |

| | | | |
|---|---|---|---|
| Treponema pallidum (*n*=3; 1.6%) | • (Meningo-) Encephalitis (*n*=2) | | • (Poly-) Neuritis cranialis (*n*=1) |
| Aspergillus (*n*=1; 0.5%) | • Encephalitis (*n*=1) | | |
| Unknown pathogen (*n*=68; 36.0%) | • (Meningo-) Encephalitis (*n*=12)<br>• Abscess (*n*=2)<br>• (Encephalo-) Myelitis (*n*=2)<br>• Myelitis + meningitis (*n*=1) | • Meningitis (*n*=41)<br>• Meningitis + (poly-) neuritis cranialis (*n*=1)<br>• Meningitis + polyradiculitis (*n*=1) | • (Poly-) Neuritis cranialis (*n*=8) |

The 189 patients with paired CSF/serum samples were divided according to their clinical syndromes and detected pathogens into three different groups: TI+: with tissue involvement, TI–: without tissue involvement, PND: peripheral nerve disease. The number of patients is given as absolute and relative values.

**Table S3. CSF NfL and GFAP raw values of different NID phenotypes paired CSF/serum samples.**

| **Demographics/ CSF parameters** | **TI+ (*n*=43)** | **TI– (*n*=59)** | **PND (*n*=47)** | ***P*-value** |
|---|---|---|---|---|
| CSF-GFAP [pg/ml] | 760 (408–1392) | 308 (205–512) | 468 (283–704) | **<0.0001**** |
| CSF-NfL [pg/ml] | 2852 (1068–7092) | 612 (389–1100) | 1356 (628–2488) | **<0.0001**** |

Group differences among TI+, TI–, and PND were assessed using the Kruskal–Wallis test** for continuous variables. Continuous data are presented as median and interquartile range (IQR, $25^{th}$–$75^{th}$ percentile). Statistical significance was set at $p<0.05$. Significant values are highlighted in bold. TI+: NID with tissue involvement, TI–: NID without tissue involvement, PND: peripheral nerve disease, CSF: cerebrospinal fluid.

**Table S4. Clinical and diagnostic characteristics in the three different groups.**

| **Characteristics** | **TI+ (*n*=48)** | **TI– (*n*=80)** | **PND (*n*=61)** | ***P*-value** |
|---|---|---|---|---|
| mRS at LP | | | | **<0.001*** |
| • 0: [+%], (n) | 0% | 0% | 0% | |
| • 1: [+%], (n) | 6.3% (3) | 73.8% (59) | 77.0% (47) | |
| • 2: [+%], (n) | 31.3% (15) | 23.8% (19) | 18.0% (11) | |
| • 3: [+%], (n) | 10.4% (5) | 1.3% (1) | 1.6% (1) | |
| • 4: [+%], (n) | 37.5% (18) | 1.3% (1) | 3.3% (2) | |
| • 5: [+%], (n) | 14.6% (7) | 0% | 0% | |
| mRS at discharge | | | | **<0.001*** |
| • 0: [+%], (n) | 43.8% (21) | 71.3% (57) | 27.9% (17) | |
| • 1: [+%], (n) | 6.3% (3) | 20.0% (16) | 49.2% (30) | |
| • 2: [+%], (n) | 16.7% (8) | 1.3% (1) | 13.1% (8) | |
| • 3: [+%], (n) | 12.5% (6) | 1.3% (1) | 1.6% (1) | |
| • 4: [+%], (n) | 8.3% (4) | 0% | 1.6% (1) | |
| • 5: [+%], (n) | 6.3% (3) | 0% | 0% | |
| • 6: [+%], (n) | 6.3% (3) | 0% | 0% | |
| New focal neurological deficits | | | | |
| • Paresis; [+%], n (+/-) | 18.8 (9/39) | 0% | 0% | **<0.001*** |
| • Aphasia; [+%], n (+/-) | 8.3% (4/44) | 0% | 0% | **<0.001*** |
| • Ataxia; [+%], n (+/-) | 12.5% (6/42) | 0% | 0% | **0.002*** |
| Further symptoms | | | | |
| • Headache; [+%], n (+/-) | 39.6% (19/29) | 98.8% (79/1) | 19.7% (12/49) | **<0.001*** |
| • Fever; [+%], n (+/-) | 25.0% (12/36) | 57.5% (46/34) | 1.6% (1/60) | **<0.001*** |

| | | | | |
|---|---|---|---|---|
| • Altered mental status/psychiatric features; [+%], n (+/-)<br>• Altered quantitative consciousness; [+%], n (+/-)<br>• Meningismus; [+%], n (+/-)<br>• Autonomic dysfunction; [+%], n (+/-) | 64.6% (31/17)<br>47.9% (23/25)<br>10.4% (5/43)<br>6.3% (3/45) | 2.5% (2/78)<br>0%<br>32.5% (26/54)<br>1.3% (1/79) | 0%<br>0%<br>0%<br>3.2% (2/59) | **<0.001*** <br>**<0.001*** <br>**<0.001*** <br>0.295* |
| New-onset seizures<br>• EEG: focal/generalized slowing; [+%], n (+/-)<br>• EEG: epileptic/status; [+%], n (+/-) | 14.6% (7/23)<br>33.3% (16/14) | 2.5% (2/8)<br>0% | 0%<br>0% | 0.213*<br>0.123* |
| Peripheral deficits<br>• Cranial nerve palsies; n (+/-), [+]<br>• Polyradiculitis; n (+/-), [+] | 8.3% (4/44)<br>2.1% (1/47) | 11.3% (9/71)<br>1.3% (1/79) | 83.6% (51/10)<br>21.3% (13/48) | **<0.001*** <br>**<0.001*** |
| MRI findings<br>• Mesiotemporal lesions; n (+/-), [+]<br>• Basal ganglia/Thalamus lesions; n (+/-), [+] | 18.8% (9/39)<br>10.4% (5/43) | 0%<br>0% | 0%<br>0% | **<0.001*** <br>**<0.001*** |

| | | | | |
|---|---|---|---|---|
| • Multifocal white matter lesions; n (+/-), [+]<br>• Brainstem lesions; n (+/-), [+]<br>• Myelitis; n (+/-), [+]<br>• Meningeal contrast-enhancement; n (+/-), [+] | 14.6% (7/41)<br>10.4% (5/43)<br>12.5% (6/42)<br>4.2% (2/46) | 0%<br>0%<br>0%<br>6.3% (5/74) | 0%<br>0%<br>0%<br>0% | **<0.001***<br>**<0.001***<br>**<0.001***<br>0.144* |
| Intrathecal IgM synthesis; [+%], n (+/-) | 37.5% (18/28) | 21.3% (17/59) | 39.3% (24/31) | **0.024*** |
| Intrathecal IgA synthesis; [+%], n (+/-) | 14.6% (7/39) | 7.5% (6/70) | 6.6% (4/51) | 0.321* |
| Intrathecal IgG synthesis; [+%], n (+/-) | 12.5% (6/40) | 3.8% (3/73) | 11.5% (7/48) | 0.122* |

We used the Pearson's $chi^2$ test* for discrete variables (mRS at LP, various new onset focal neurological deficits, various further symptoms, various peripheral deficits, various MRI findings) to determine differences between the three groups. The values for discrete variables are given as absolute and relative counts. *P*-values <0.05 were considered statistically significant. Significant values are highlighted in bold. TI+: with tissue involvement, TI–: without tissue involvement, PND: peripheral nerve disease, mRS: modified Rankin scale, EEG: electroencephalogram, MRI: magnetic resonance imaging.

**Table S5 CSF findings in the different diseases, separated by pathogens.**

| **CSF parameters** | **TBEV (*n*=12)** | **Staph./Strep./ Citro. (*n*=4)** | **TP (*n*=3)** | **ASP (*n*=1)** | **EBV (*n*=1)** | **JCV (*n*=1)** | **HepE (*n*=1)** | **HSV-1/-2 (*n*=11)** | **VZV (*n*=50)** | **BB (*n*=37)** | **UP (*n*=68)** | ***P*-value** |
|---|---|---|---|---|---|---|---|---|---|---|---|---|
| Leucocyte count [/µl] | 55.5 (29.75-94) | 330.0 (1156.25-142.75) | 3.0 (1.0-3.0) | 42.0 | 17.0 | 1.0 | 18.0 | 77.0 (19.0-164.0) | 112.0 (49.25-314.75) | 150.0 (80.0-239.0) | 120.0 (35.25-219.75) | **0.002**** |
| Lactate [mmol/l] | 1.8 (1.71-2.08) | 3.12 (2.39-4.63) | 1.6 (1.41-1.6) | 3.2 | 1.44 | 1.51 | 1.7 | 2.1 (1.8-2.6) | 2.1 (1.7-2.7) | 2.3 (1.87-2.7) | 2.0 (1.7-2.5) | **0.019**** |
| Total protein [mg/l] | 773.0 (587.75-1116.0) | 707.0 (576.5-1753.25) | 4020 (238.0-402.0) | 1087.0 | 497.0 | 400.0 | 956.0 | 664.0 (468.0-1167.0) | 792.0 (542.75-1365.0) | 1676.0 (1143.0-2154.0) | 691.0 (452.25-958.75) | **<0.001**** |
| QAlb (CSF/S) | 10.7 (8.85-19.2) | 11.35 (8.68-31.95) | 4.5 (2.8-4.5) | 18.1 | 7.2 | 6.7 | 15.2 | 8.97 (6.8-19.6) | 12.2 (7.65-23.75) | 21.8 (12.4-32.7) | 9.3 (6.18-14.33) | **<0.001**** |
| CSF-CXCL13 [pg/ml] | 24.0 (5.9-62.0) | 212.0 (4.0-212.0) | 189.0 (19.0-189.0) | 480.0 | - | - | - | 222.5 (21.25-368.0) | 78.0 (26.0-179.0) | 6150.0 (1036.0-14455.0) | 63.0 (20.0-232.75) | **<0.001**** |
| CSF-specific oligoclonal IgG bands; [+%], n (+/-) | 41.7% (5/7) | 0% (0/4) | 100% (3/0) | 0% (0/1) | 100% (1/0) | 0% (0/1) | 0% (0/1) | 36.4% (4/11) | 32.0% (16/50) | 86.4% (32/5) | 23.5% (16/66) | **<0.001*** |
| Intrathecal IgM synthesis; [+%], n (+/-) | 66.7% (8/4) | 50% 82/2) | 33.3% (1/2) | 0% (0/1) | 0% (0/1) | 0% (0/1) | 0% (0/1) | 9.1% (1/11) | 12.0% (6/37) | 83.8% (31/5) | 14.7% (10/54) | **<0.001*** |

| | | | | | | | | | | | | |
|---|---|---|---|---|---|---|---|---|---|---|---|---|
| Intrathecal IgA synthesis; [+%], n (+/-) | 0% (0/12) | 0% (0/4) | 0% (0/3) | 0% (0/1) | 100% (1/0) | 0% (0/1) | 0% (0/1) | 0% (0/11) | 2.0% (1/42) | 18.9% (7/29) | 11.8% (8/56) | **0.028*** |
| Intrathecal IgG synthesis; [+%], n (+/-) | 8.3% (1/11) | 0% (0/4) | 0% (0/3) | 0% (0/1) | 0% (0/1) | 0% (0/1) | 0% (0/1) | 0% (0/11) | 0% (0/43) | 24.3% (9/27) | 4.4% (3/61) | **<0.001*** |

For determining the differences between the various pathogens, the Pearson's chi$^2$ test* was used for discrete variables (CSF-specific oligoclonal IgG bands, intrathecal IgM/IgA/IgG synthesis) and the Kruskal Wallis test** for continuous variables (leucocyte count, lactate, total protein, QAlb, CSF-CXCL13). The values for discrete variables are given as absolute and relative counts. The values for continuous variables are given as median and interquartile range (IQR; 25$^{th}$–75$^{th}$ percentile). For *p*-values <0.05, we assumed statistical significance. Significant values are highlighted in bold. CSF: cerebrospinal fluid, QAlb: CSF-albumin/serum-albumin, CXCL13: C-X-C-motif chemokine ligand 13, IgM/IgA/IgG: immunoglobulin M/A/G, TBEV: tick-borne encephalitis virus, Staph./Strep./ Citro.: Staphylococcus aureus/epidermidis/Streptococcus anginosus/Citrobacter freundii, TP: treponema pallidum, ASP: aspergillus, EBV: Epstein-Barr virus, JCV: John Cunningham virus, HepE: hepatitis E virus, HSV-1/-2: herpes simplex virus type 1/2, VZV; varicella-zoster virus, BB: borrelia burgdorferi, UP: unknown pathogen.

**Table S6. Correlation assessment between CSF and blood biomarkers and clinical data (Part 1).**

| | sGFAP vs. | | | | sNfL vs. | | | | CSF-GFAP vs. | | | | CSF-NfL vs. | | | |
|---|---|---|---|---|---|---|---|---|---|---|---|---|---|---|---|---|
| | CSF-CXCL13 | CSF-lactate | QAlb | Duration of hospitalization [days] | CSF-CXCL13 | CSF-lactate | QAlb | Duration of hospitalization [days] | CSF-CXCL13 | CSF-lactate | QAlb | Duration of hospitalization [days] | CSF-CXCL13 | CSF-lactate | QAlb | Duration of hospitalization [days] |
| **Total neuroinfectious cohort** | | | | | | | | | | | | | | | | |
| **Spearman's *r*** | 0.15 | 0.17 | 0.11 | 0.26 | 0.31 | 0.15 | 0.29 | 0.26 | 0.21 | 0.35 | 0.44 | 0.31 | 0.26 | 0.32 | 0.47 | 0.35 |
| **95% confidence interval of *r*** | -0.05 – 0.33 | 0.03 – 0.31 | -0.04 –0.26 | 0.12 – 0.39 | 0.12 – 0.48 | -0.002 – 0.29 | 0.15 –0.42 | 0.11 – 0.39 | -0.01 – 0.41 | 0.19 – 0.48 | 0.30 – 0.57 | 0.15 – 0.45 | 0.04 –0.45 | 0.16 – 0.46 | 0.33 – 0.59 | 0.19 – 0.49 |
| ***P*-value** | 0.1366 | 0.0177 | 0.1322 | 0.0003 | 0.0013 | 0.0467 | <0.0001 | 0.0004 | 0.0537 | <0.0001 | <0.0001 | 0.0002 | 0.0193 | <0.0001 | <0.0001 | <0.0001 |
| ***P*-value summary** | ns | * | ns | *** | ** | * | **** | *** | ns | **** | **** | *** | * | **** | **** | **** |
| ***n*** | 105 | 185 | 184 | 186 | 105 | 185 | 184 | 186 | 84 | 147 | 147 | 148 | 84 | 147 | 147 | 148 |
| **TI+** | | | | | | | | | | | | | | | | |
| **Spearman's *r*** | 0.29 | 0.29 | 0.02 | 0.11 | 0.67 | 0.21 | 0.30 | 0.13 | 0.26 | 0.45 | 0.31 | 0.07 | 0.50 | 0.32 | 0.38 | 0.21 |
| **95% confidence interval of *r*** | -0.10 – 0.60 | -0.005 – 0.54 | -0.28 – 0.31 | -0.19 – 0.40 | 0.39 – 0.84 | -0.09 – 0.48 | 0.01 – 0.55 | -0.17 – 0.42 | -0.16 – 0.60 | 0.17 – 0.67 | 0.003 – 0.56 | -0.25 – 0.37 | 0.12 – 0.75 | 0.02 –0.57 | 0.09 – 0.62 | -0.11 – 0.49 |
| ***P*-value** | 0.1284 | 0.0473 | 0.9204 | 0.4574 | <0.0001 | 0.1464 | 0.0381 | 0.3728 | 0.2031 | 0.0022 | 0.0421 | 0.6768 | 0.0113 | 0.0346 | 0.0110 | 0.1809 |
| ***P*-value summary** | ns | * | ns | ns | **** | ns | * | ns | ns | ** | * | ns | * | * | * | ns |
| ***n*** | 29 | 47 | 47 | 46 | 29 | 47 | 47 | 46 | 25 | 43 | 43 | 42 | 25 | 43 | 43 | 42 |
| **TI–** | | | | | | | | | | | | | | | | |
| **Spearman's *r*** | 0.21 | 0.07 | -0.007 | -0.014 | 0.14 | -0.04 | 0.09 | 0.04 | 0.33 | 0.25 | 0.34 | 0.22 | -0.005 | 0.11 | 0.32 | 0.27 |
| **95% confidence interval of *r*** | -0.14 – 0.51 | -0.16 – 0.29 | -0.23 – 0.22 | -0.24 – 0.21 | -0.21 – 0.46 | -0.27 – 0.19 | -0.14 – 0.31 | -0.19 – 0.26 | -0.06 – 0.64 | -0.02 – 0.48 | 0.08 – 0.55 | -0.04 – 0.46 | -0.40 – 0.39 | -0.16 – 0.36 | 0.06 – 0.54 | 0.01 – 0.50 |
| ***P*-value** | 0.2232 | 0.5351 | 0.9494 | 0.9032 | 0.4169 | 0.7079 | 0.4494 | 0.7343 | 0.0890 | 0.0630 | 0.0085 | 0.0934 | 0.9795 | 0.4156 | 0.0132 | 0.0348 |
| ***P*-value summary** | ns | ns | ns | ns | ns | ns | ns | ns | ns | ns | ** | ns | ns | ns | * | * |
| ***n*** | 36 | 79 | 79 | 80 | 36 | 79 | 79 | 80 | 27 | 58 | 59 | 59 | 27 | 58 | 59 | 59 |
| **PND** | | | | | | | | | | | | | | | | |
| **Spearman's *r*** | 0.32 | 0.17 | 0.16 | 0.37 | 0.16 | 0.39 | 0.35 | 0.45 | 0.21 | 0.39 | 0.56 | 0.25 | 0.32 | 0.62 | 0.73 | 0.26 |
| **95% confidence interval of *r*** | 0.006 – 0.59 | -0.093 – 0.42 | -0.11 – 0.40 | 0.12 – 0.57 | -0.17 – 0.45 | 0.14 – 0.59 | 0.09 – 0.56 | 0.21 – 0.64 | -0.16 – 0.53 | 0.11 – 0.62 | 0.31 – 0.74 | -0.05 – 0.51 | -0.04 – 0.61 | 0.40 – 0.78 | 0.55 – 0.85 | -0.03 – 0.52 |
| ***P*-value** | 0.0405 | 0.1857 | 0.2429 | 0.0039 | 0.3351 | 0.0024 | 0.0072 | 0.0003 | 0.2399 | 0.0066 | <0.0001 | 0.0934 | 0.0763 | <0.0001 | <0.0001 | 0.0713 |
| ***P*-value summary** | * | ns | ns | ** | ns | ** | ** | *** | ns | ** | **** | ns | ns | **** | **** | ns |
| ***n*** | 40 | 59 | 58 | 60 | 40 | 59 | 58 | 60 | 32 | 46 | 45 | 47 | 32 | 46 | 45 | 47 |

mRS: modified Rankin Scale, CSF: cerebrospinal fluid, QAlb: CSF/serum albumin ratio, CXCL13: C-X-C-motif chemokine ligand 13, ns: not significant, TI+/TI–: with/without tissue involvement, PND: peripheral nerve disease, LP: lumbar puncture.

**Table S7. Correlation assessment between CSF and blood biomarkers and clinical data (Part 2).**

| | sGFAP vs. | | | | sNfL vs. | | | | CSF-GFAP vs. | | | | CSF-NfL vs. | | | |
|---|---|---|---|---|---|---|---|---|---|---|---|---|---|---|---|---|
| | mRS at LP | mRS at discharge | Leucocyte count | Time from onset to LP [days] | mRS at LP | mRS at discharge | Leucocyte count | Time from onset to LP [days] | mRS at LP | mRS at discharge | Leucocyte count | Time from onset to LP [days] | mRS at LP | mRS at discharge | Leucocyte count | Time from onset to LP [days] |
| **Total neuroinfectious cohort** | | | | | | | | | | | | | | | | |
| **Spearman's *r*** | 0.34 | 0.23 | -0.28 | 0.31 | 0.32 | 0.30 | -0.31 | 0.33 | 0.38 | 0.26 | -0.11 | 0.26 | 0.45 | 0.31 | -0.19 | 0.41 |
| **95% confidence interval of *r*** | 0.20 – 0.46 | 0.08 – 0.36 | -0.41 – - 0.14 | 0.18 – 0.44 | 0.18 – 0.45 | 0.15 – 0.43 | -0.43– - 0.16 | 0.19 – 0.45 | 0.23 – 0.51 | 0.1 – 0.41 | -0.27 – 0.062 | 0.09 – 0.41 | 0.31 – 0.57 | 0.14 – 0.45 | -0.35 – -0.03 | 0.26 – 0.54 |
| ***P*-value** | <0.0001 | 0.0028 | <0.0001 | <0.0001 | <0.0001 | <0.0001 | <0.0001 | <0.0001 | <0.0001 | 0.0018 | 0.2031 | 0.0015 | <0.0001 | 0.0003 | 0.0174 | <0.0001 |
| ***P*-value summary** | **** | ** | **** | **** | **** | **** | **** | **** | **** | ** | ns | ** | **** | *** | * | **** |
| ***n*** | 187 | 175 | 187 | 187 | 187 | 175 | 187 | 187 | 149 | 140 | 149 | 149 | 149 | 140 | 149 | 149 |
| **TI+** | | | | | | | | | | | | | | | | |
| **Spearman's *r*** | 0.23 | 0.33 | -0.22 | -0.007 | 0.31 | 0.32 | -0.24 | 0.07 | 0.30 | 0.37 | 0.002 | 0.21 | 0.27 | 0.29 | -0.18 | 0.25 |
| **95% confidence interval of *r*** | -0.07 – 0.49 | 0.03 – 0.58 | -0.49 – 0.08 | -0.30 – 0.29 | 0.02 – 0.56 | 0.01 – 0.57 | -0.50 – 0.06 | -0.22 – 0.36 | -0.01 – 0.55 | 0.05 – 0.61 | -0.31 – 0.31 | -0.11 – 0.49 | -0.04 – 0.54 | -0.03 – 0.56 | -0.46 – 0.14 | -0.07 – 0.51 |
| ***P*-value** | 0.1271 | 0.0269 | 0.1332 | 0.9597 | 0.0320 | 0.0346 | 0.1082 | 0.6342 | 0.0545 | 0.0203 | 0.9875 | 0.1794 | 0.0761 | 0.0705 | 0.2530 | 0.1123 |
| ***P*-value summary** | ns | * | ns | ns | * | * | ns | ns | ns | * | ns | ns | ns | ns | ns | ns |
| ***n*** | 47 | 44 | 47 | 47 | 47 | 44 | 47 | 47 | 43 | 40 | 43 | 43 | 43 | 40 | 43 | 43 |
| **TI–** | | | | | | | | | | | | | | | | |
| **Spearman's *r*** | -0.04 | 0.02 | -0.32 | 0.37 | 0.07 | 0.01 | -0.28 | 0.40 | -0.10 | -0.04 | 0.003 | 0.19 | 0.10 | 0.16 | -0.12 | 0.48 |
| **95% confidence interval of *r*** | -0.26 – 0.19 | -0.21 – 0.25 | -0.51 – - 0.09 | 0.15 –0.55 | -0.16 – 0.29 | -0.22 – 0.24 | -0.47 – - 0.06 | 0.19 –0.57 | -0.36 – 0.16 | -0.31 – 0.23 | -0.26 – 0.27 | -0.08 – 0.43 | -0.17 – 0.35 | -0.12 – 0.41 | -0.37 – 0.15 | 0.25 –0.66 |
| ***P*-value** | 0.7250 | 0.8691 | 0.0041 | 0.0008 | 0.5216 | 0.9248 | 0.0125 | 0.0002 | 0.4381 | 0.7711 | 0.9841 | 0.1499 | 0.4471 | 0.2429 | 0.3786 | 0.0001 |
| ***P*-value summary** | ns | ns | ** | *** | ns | ns | * | *** | ns | ns | ns | ns | ns | ns | ns | *** |
| ***n*** | 80 | 75 | 80 | 80 | 80 | 75 | 80 | 80 | 59 | 56 | 59 | 59 | 59 | 56 | 59 | 59 |
| **PND** | | | | | | | | | | | | | | | | |
| **Spearman's *r*** | 0.20 | 0.10 | -0.09 | 0.39 | 0.29 | 0.11 | 0.05 | 0.30 | 0.40 | 0.18 | 0.05 | 0.25 | 0.53 | 0.23 | 0.19 | 0.37 |
| **95% confidence interval of *r*** | -0.07 – 0.44 | -0.18 – 0.36 | -0.34 – 0.18 | 0.15 –0.59 | 0.03 – 0.51 | -0.17 – 0.37 | -0.21 – 0.31 | 0.04 –0.52 | 0.12 – 0.62 | -0.13 – 0.46 | -0.25 – 0.34 | -0.04 – 0.51 | 0.28 – 0.72 | -0.08 – 0.50 | -0.11 – 0.46 | 0.08 –0.60 |
| ***P*-value** | 0.1303 | 0.4874 | 0.5002 | 0.0019 | 0.0242 | 0.4262 | 0.7136 | 0.0207 | 0.0049 | 0.2333 | 0.7476 | 0.0861 | 0.0001 | 0.1273 | 0.1918 | 0.0117 |
| ***P*-value summary** | ns | ns | ns | ** | * | ns | ns | * | ** | ns | ns | ns | *** | ns | ns | * |
| ***n*** | 60 | 56 | 60 | 60 | 60 | 56 | 60 | 60 | 47 | 44 | 47 | 47 | 47 | 44 | 47 | 47 |

mRS: modified Rankin Scale, CSF: cerebrospinal fluid, TI+/TI–: with/without tissue involvement, PND: peripheral nerve disease, LP: lumbar puncture.

### Supplementary Methods

*Study design, participants and selection process*

Patients undergoing diagnostic lumbar puncture (LP) at the University Hospital of Ulm, Germany, from August 2009 to November 2023, for suspected meningitis, encephalitis, cranial nerve palsies, or polyradiculitis who consented to scientific use of their biosamples were retrospectively included in this monocentric cohort study (*n*=451). Medical records, including CSF and blood findings, were reviewed for case selection. Only patients with stored CSF and/or serum obtained before initiation of anti-infective therapy were eligible, leading to exclusion of 141 patients; a further 66 were excluded due to insufficient sample volume for NfL or GFAP measurement, and 53 showed no infectious CNS inflammation. According to predefined criteria, 189 patients were included (serum: *n*=187; CSF: *n*=149). Patients were classified as having encephalitis, meningitis, myelitis, cranial nerve palsies, polyradiculitis, or combinations thereof based on clinical records and published case definitions (Table S1; Supplementary Material) [1-5]. Pathogens were identified by polymerase chain reaction (PCR) and/or pathogen-specific CSF/serum antibody indices or by cultural/microscopic evidence; neurosyphilis was additionally confirmed using the Venereal Disease Research Laboratory (VDRL) test (CSF), the fluorescent treponema antibody absorption (FTA-ABS) test (serum), and a Treponema pallidum particle agglutination/haemagglutination (TPPA/TPHA) test (CSF/serum) [6].

Detected pathogens included TBEV (*n*=12), HSV-1/-2 (*n*=11), VZV (*n*=50), Epstein-Barr virus (EBV; *n*=1), hepatitis E virus (HepE; *n*=1), John Cunningham virus (JCV; *n*=1), bacterial or fungal pathogens (staphylococcus aureus/epidermidis/streptococcus anginosus/citrobacter freundii, aspergillus; *n*=5), Borrelia burgdorferi (*n*=37), and Treponema pallidum (*n*=3); no pathogen was identified in 36.0% (*n*=68), although CSF, blood, and clinical findings met case definitions consistent with CNS infection (Table S1; Supplementary Material). Patients were further grouped by syndrome (Table S2;

Supplementary Material) into NID with tissue involvement (TI+; (meningo-)encephalitis, (encephalo-)myelitis, abscess), NID without tissue involvement (TI–; meningitis), and peripheral nerve disease (PND; (poly-)neuritis cranialis, polyradiculitis), with mixed presentations assigned according to the predominant syndrome. All patients underwent cranial imaging, mainly magnetic resonance imaging (MRI), before LP; electroencephalogram (EEG) was performed when clinically indicated. Outcome was assessed using the modified Rankin Scale (mRS) at LP and discharge. Among 189 patients, 57 had a second (TI+: 10/TI–: 33/PND: 14), 7 a third (TI+: 1/TI–: 3/PND: 3), and 2 a fourth (TI+: 1/TI–: 1) LP within 30 days.

A patient control group (PC) comprised 116 patients (76 with paired CSF/serum samples and 40 with serum only) and no evidence of inflammatory CNS disease (CSF cell count <5/µl, lactate <2.6 mmol/l, no intrathecal immunoglobulin synthesis, and no/mild blood–CSF barrier dysfunction). Clinical diagnoses mainly included tension-type headache (*n*=36), migraine (*n*=14), parainfectious headache (*n*=2), idiopathic intracranial hypertension (*n*=2), somatoform disorders (*n*=30), non-infectious cranial nerve palsies (*n*=7), transient global amnesia (*n*=7), benign paroxysmal positional vertigo (*n*=4), dizziness (*n*=3), depression (*n*=3), anxiety disorder (*n*=1), atypical facial pain (*n*=2), syncope (*n*=4), and ocular myasthenia gravis (*n*=1).

*CSF analyses*

Upon diagnostic lumbar- and venipuncture, white blood cell count (WBCC) was determined manually using the Fuchs-Rosenthal chamber (normal ≤4 leucocytes/µl). CSF-lactate was measured using a photometer (normal: 1.7-2.6 mmol/l, AU400/AU680 Clinical Chemistry Analyzers (Olympus/Beckman Coulter, Krefeld, Germany)). Total protein (g/l), albumin (mg/l), and immunoglobulins A, M, and G (mg/l) in CSF and serum were detected nephelometrically (BN Prospec nephelometer (Siemens, Munich,

Germany)). For defining the blood-CSF barrier function, we used the CSF/serum albumin ratio (QAlb) with the age-related cut-off for Qlim(Alb)=(4+age/15)×$10^{-03}$ [7]. Disturbance of the blood-CSF barrier was assumed for QAlb>Qlim(Alb).[7] QAlb was also used for index calculations of NfL and GFAP: CSF/serum ratio of each biomarker divided by QAlb. Oligoclonal IgG bands (OCB) in CSF and serum were assessed by isoelectric focusing (IEF). To calculate their quantitative expression, we built CSF/serum ratios of the immunoglobulins (QIgG/QIgA/QIgM) to calculate their quantitative expression. Using QAlb, their upper limits were calculated with Reiber's formula for the hyperbolic function [8]. For QIg>Qlim(Ig), intrathecal synthesis was assumed, and their intrathecal fraction was determined [8]. Measurements for specific IgM- and IgG- antibodies against HSV-1, VZV, EBV, Borrelia burgdorferi, and TBEV were performed using an enzyme-linked immunosorbent assay (ELISA; Gold Standard Diagnostics Europe (former Genzyme Virotech), Rüsselsheim, Germany). For calculation of their intrathecal synthesis, we used the following formula: specific antibody index (AI)=QIg(spec)/QIg(total) for QIg(total)<Qlim(Ig) and QIg(spec)/Qlim(Ig) for QIg(total)>Qlim(Ig) [9]. Antibody indices ≥1.5 indicated an intrathecal synthesis of the tested antigen [9].

*Quantification of GFAP and NfL in CSF and serum*

Serum samples were diluted 1:2, while CSF samples were diluted 1:4 prior to analysis. Each assay run included two quality control samples measured in duplicate, with inter-assay variability maintained below 20%. For GFAP, the Simple Plex Human GFAP ($2^{nd}$ Gen) Assay Cartridge (SPCKB-PS-009134) was employed, featuring a dynamic range of 2.5-9600 pg/mL. NfL quantification utilized the Simple Plex Human NF-L Cartridge (SPCKB-PS-002448), with a dynamic range of 2.7-10290 pg/mL. All measurements were conducted on aliquots subjected to an identical number of freeze–thaw cycles to ensure consistency and reliability across samples, and assays were performed according to the manufacturer's instructions.
